# Expression of mitochondrial oxidative stress response genes in muscle is associated with mitochondrial respiration, physical performance, and muscle mass in the Study of Muscle, Mobility and Aging (SOMMA)

**DOI:** 10.1101/2023.11.05.23298108

**Authors:** Gregory J Tranah, Haley N Barnes, Peggy M Cawthon, Paul M Coen, Karyn A Esser, Russell T Hepple, Zhiguang Huo, Philip A Kramer, Frederico G. S. Toledo, Daniel S Evans, Steven R Cummings

## Abstract

Gene expression in skeletal muscle of older individuals may reflect compensatory adaptations in response to oxidative damage that preserve tissue integrity and maintain function. Identifying associations between oxidative stress response gene expression patterns and mitochondrial function, physical performance, and muscle mass in older individuals would further our knowledge of mechanisms related to managing molecular damage that may be targeted to preserve physical resilience. To characterize expression patterns of genes responsible for the oxidative stress response, RNA was extracted and sequenced from skeletal muscle biopsies collected from 575 participants (≥70 years old) from the Study of Muscle, Mobility and Aging. Expression levels of twenty-one protein coding RNAs related to the oxidative stress response were analyzed in relation to six phenotypic measures, including: maximal mitochondrial respiration from muscle biopsies (Max OXPHOS), physical performance (VO_2_ peak, 400m walking speed, and leg strength), and muscle size (thigh muscle volume and whole-body D3Cr muscle mass). The mRNA level of the oxidative stress response genes most consistently associated across outcomes are preferentially expressed within the mitochondria. Higher expression of mRNAs that encode generally mitochondria located proteins *SOD2*, *TRX2*, *PRX3*, *PRX5*, and *GRX2* were associated with higher levels of mitochondrial respiration and VO_2_ peak. In addition, greater *SOD2, PRX3,* and *GRX2* expression was associated with higher physical performance and muscle size. Identifying specific mechanisms associated with high functioning across multiple performance and physical domains may lead to targeted antioxidant interventions with greater impacts on mobility and independence.

## 1. INTRODUCTION

Mobility often declines with age, often leading to major mobility disability and subsequently decreased quality of life. Mobility disability leads to greater difficulty with activities of daily living, loss of independence, institutionalization, and greater annual health care costs for affected persons (Bouchard et al., 1999; Hardy, Kang, Studenski, & Degenholtz, 2011; Penninx et al., 2004; Studenski et al., 2011). The only proven intervention, exercise, modestly reduces the risk of developing mobility disability (Pahor et al., 2014), highlighting the need for new intervention targets. However, the age-related changes in muscle responsible for major mobility disability are poorly understood. Oxidative stress is believed to be a major age-related cause of declining muscle mass, function, and mitochondrial energetics, which are critical for mobility. The antioxidant response elements (AREs) are known to increase the expression of antioxidants to scavenge damaging oxidants and improve oxidative stress recovery (Raghunath et al., 2018). However, conflicting reports suggests antioxidant signaling in muscle of older individuals may be impaired (Zhang, Davies, & Forman, 2015), disrupting antioxidant expression and oxidative stress recovery, and blunting the effects of exercise. Very few human studies have obtained the muscle biopsies required to assess biological mechanisms responsible for the age-related loss of mobility, fitness, and strength (Butikofer, Zurlinden, Bolliger, Kunz, & Sonderegger, 2011; Carnio et al., 2014; Jang et al., 2010; Kulakowski, Parker, & Personius, 2011; Nicklas et al., 2008; Tyrrell et al., 2015). This lack of knowledge is a major obstacle to developing new interventions for our rapidly aging society.

Mitochondrial dysfunction and its attendant increase in oxidative stress is an important component in the loss of muscle mass, increase in muscle damage, and reduction in muscle contractility that are characteristic of sarcopenia (Michelucci, Liang, Protasi, & Dirksen, 2021). Oxidative phosphorylation (OXPHOS) requires 4-electron reduction of oxygen to water to establish the proton-motive force required to phosphorylate ADP into ATP to meet muscle energy demands (Mailloux, 2018; Napolitano, Fasciolo, & Venditti, 2021). Reactive oxygen species (ROS), such as superoxide and hydrogen peroxide are produced via multiple sites in the mitochondria and other sources via one and two electron reduction of oxygen (Mailloux, 2018; Napolitano et al., 2021). ROS can directly damage muscle proteins, lipids, and DNA in a reversible or irreversible manner, leading to mitochondrial and contractile dysfunction and cell death (Milkovic, Cipak Gasparovic, Cindric, Mouthuy, & Zarkovic, 2019). Antioxidant defenses to ROS are not only enriched in the mitochondria to directly degrade ROS formed by nutrient oxidation and respiration but are also critical to quenching extramitochondrial ROS. These primary antioxidant defenses include several superoxide dismutases, glutathione peroxidases, peroxiredoxins, thioredoxins, glutaredoxins, and catalase (Mailloux, 2018; Napolitano et al., 2021; Powers & Jackson, 2008). Oxidative damage to contractile proteins increases with aging and a few small studies have correlated damage with compromised muscle function (Andersson et al., 2011; Barreiro, 2016; Barreiro et al., 2006; Baumann, Kwak, Liu, & Thompson, 2016; Choksi & Papaconstantinou, 2008; Gianni, Jan, Douglas, Stuart, & Tarnopolsky, 2004; Pansarasa et al., 2000; Snow, Fugere, & Thompson, 2007; Thompson, Durand, Fugere, & Ferrington, 2006). Contractile proteins, myosin thick filaments, and actin containing tropomyosin and troponin thin filaments are especially susceptible to oxidation (Fedorova, Kuleva, & Hoffmann, 2010). While the mitochondrial and extramitochondrial antioxidant enzymes are well characterized with regard to specific roles and cellular compartmentalization, precise molecular mechanisms connecting oxidative stress and the responses to this stress to mobility decline are not fully understood.

To address these gaps, we performed gene expression in skeletal muscle biopsies collected as part of the Study of Muscle, Mobility and Aging (SOMMA) (Cummings et al., 2023). SOMMA is a large, longitudinal study of older people designed to understand the contributions of skeletal muscle mass and key properties of muscle tissue from biopsies to mobility. In this investigation, we examined associations between expression levels of twenty-one canonical mitochondrial and extramitochondrial antioxidant genes and muscle mitochondrial function. Since mitochondrial function is posited to be a key biologic driver of age-related deficits (Lopez-Otin, Blasco, Partridge, Serrano, & Kroemer, 2013), we also hypothesized that expression of antioxidant genes would be associated with 400m walking speed, VO_2_ peak, leg strength, thigh muscle volume and whole-body muscle mass.

## 2. RESULTS

### 2.1 Participant characteristics

A total of 879 participants provided consent and completed baseline measurements across both clinical sites (Figure 1). Of the 879 participants with complete baseline measures, 591 participants had RNA sequencing completed and 575 of these had high quality sequencing and complete covariate data. The characteristics of the study population with complete RNA sequencing and complete covariate data are presented in Table 1.

**Figure 1.**
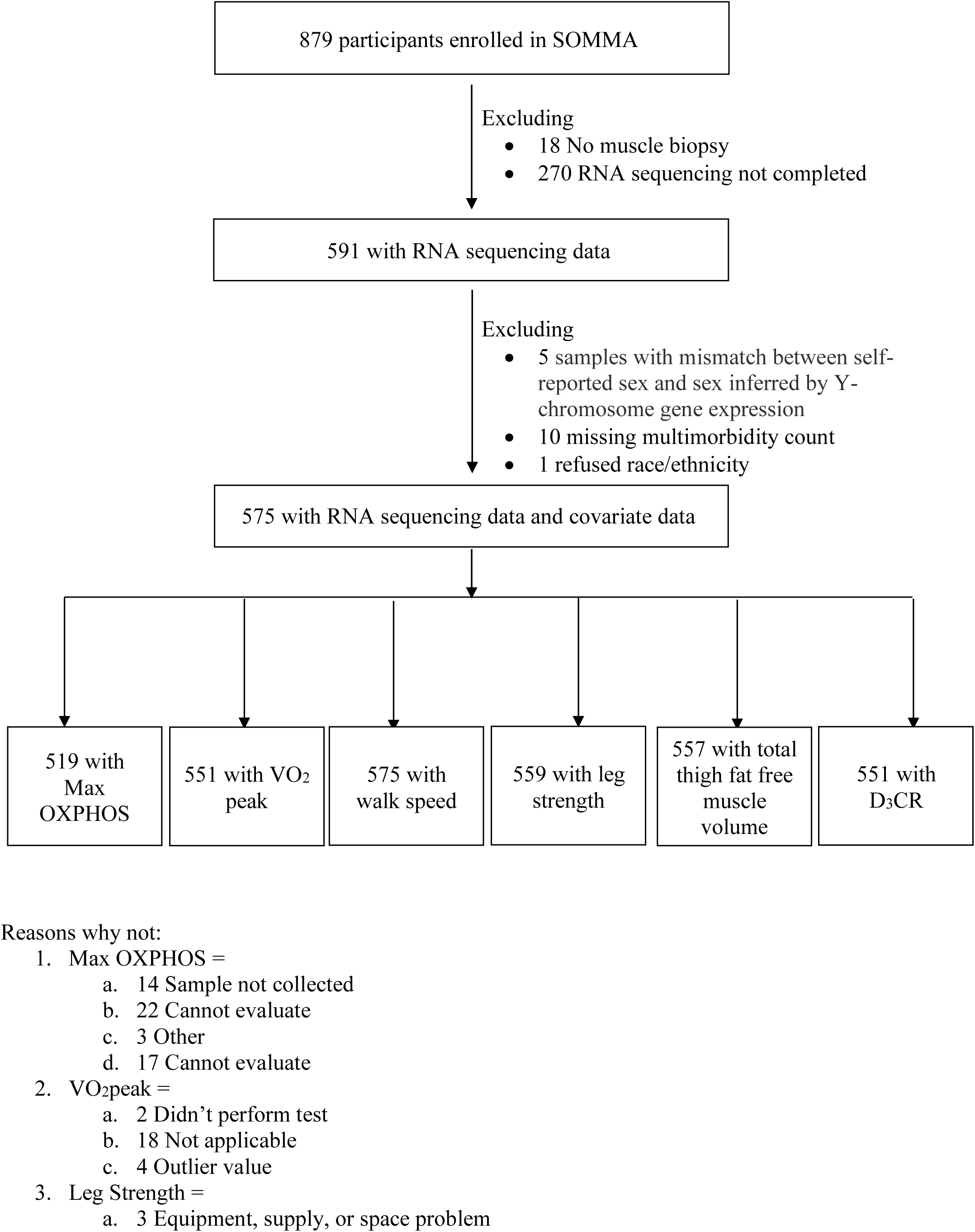
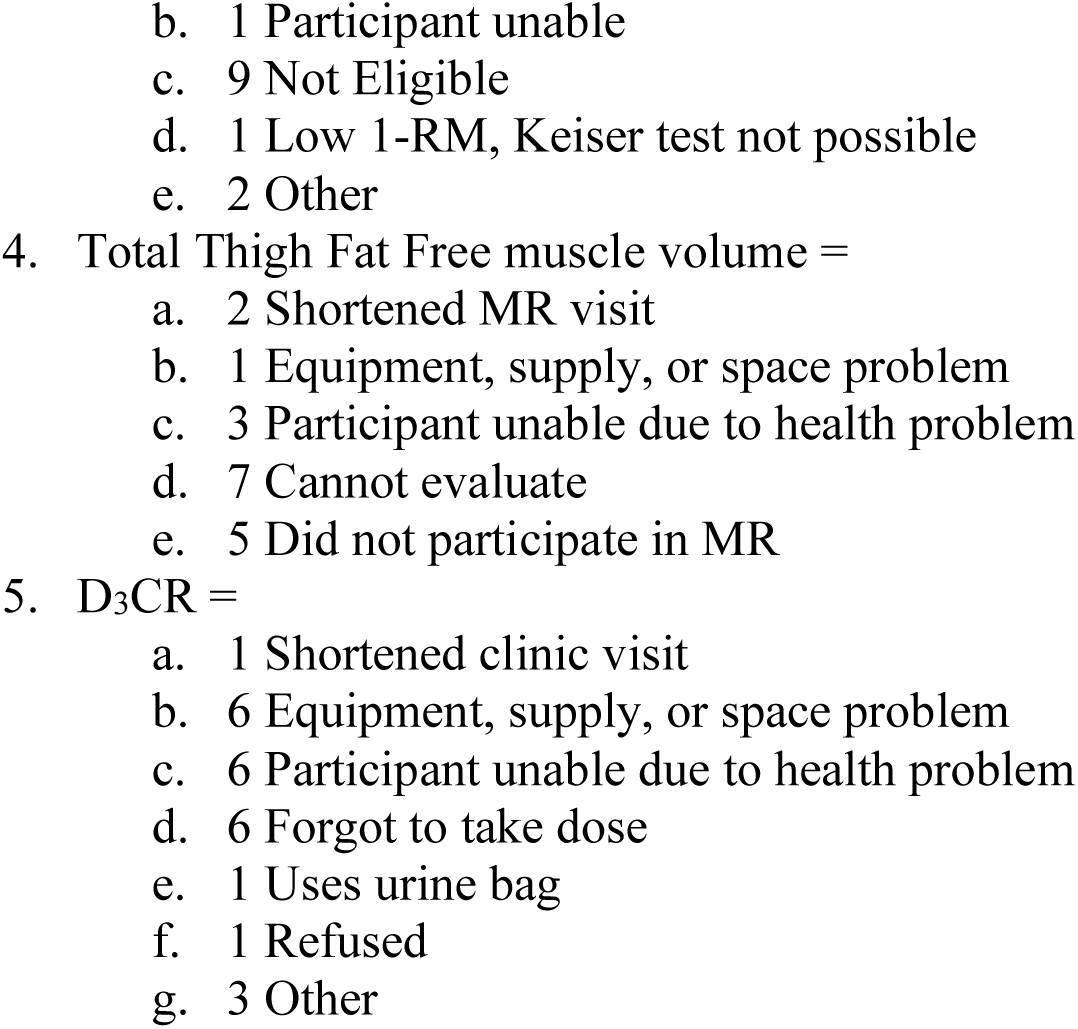
Participant inclusions. SOMMA participants selected for cross-sectional analyses.

**Table 1.**
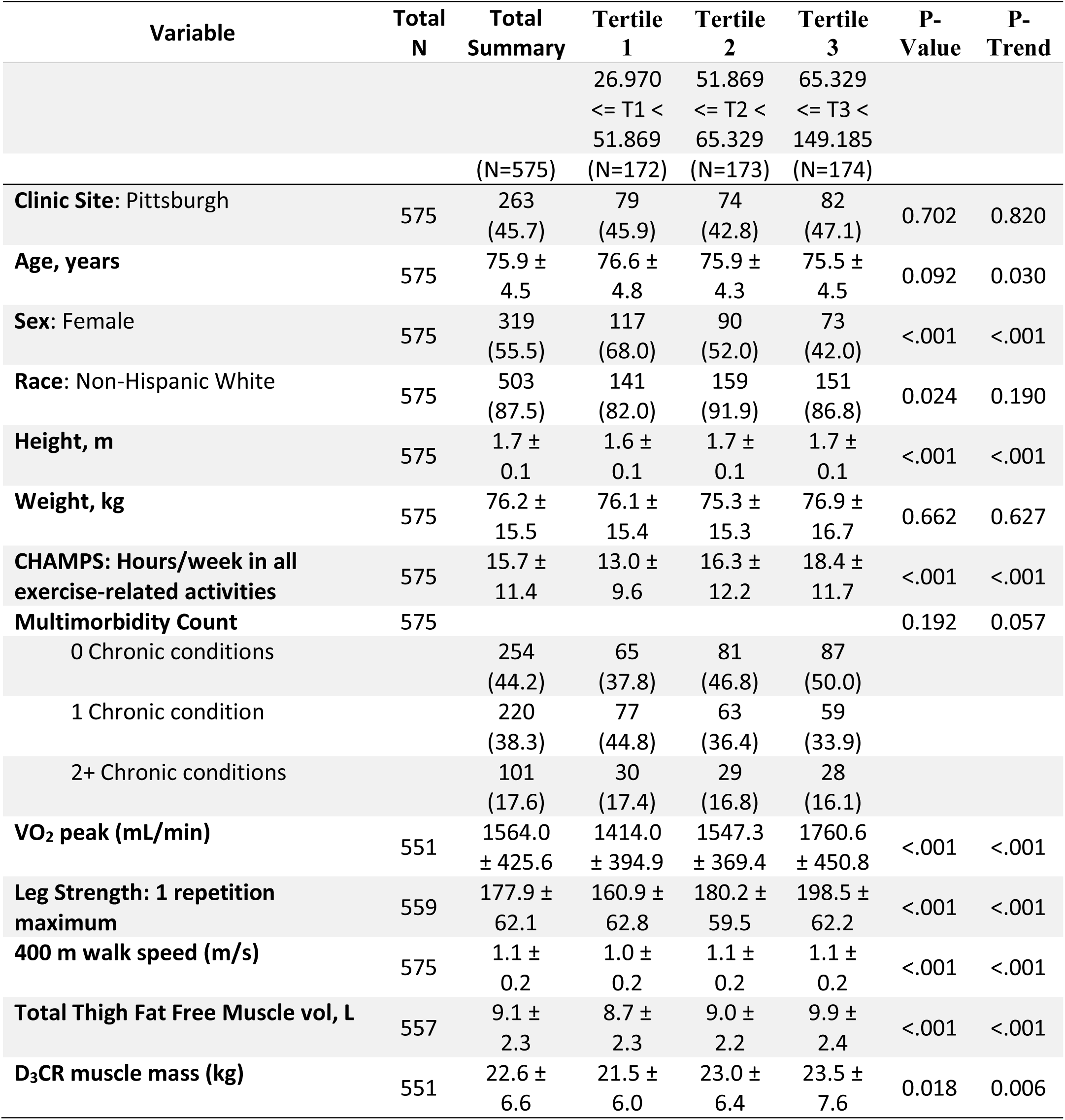
Baseline characteristics of included SOMMA participants also stratified by tertiles of Max OXPHOS. Data shown as n (%), mean ± Standard Deviation. P-values are presented for variables across tertiles of P1-Max OXPHOS. P-values for continuous variables from ANOVA for normally distributed data, a Kruskal-Wallis test for skewed data. P for linear trend across categories was calculated with linear regression models for those normally distributed variables, a Jonckheere-Terpstra test for skewed data. P-values for categorical data from a chi-square test for homogeneity. P for trend was calculated with the Jonckheere-Terpstra test.

### 2.2 RNA (human Ensembl genes (ENSG)) detection

The mean, median, and SD of the PCR duplicate percent per sample was 59%, 56% and 9%, respectively (Supplementary Table 2). After PCR duplicates were removed, the number of aligned reads per sample was high (mean=69,117,209, median = 71,313,059, SD = 14,444,848, range = 12,853,785-102,724,183).

### 2.3 Association of Max OXPHOS with gene expression

All results report log base 2-fold changes reflecting the change in gene expression per one SD unit increase in each trait. Of the twenty-one oxidative stress response genes analyzed for associations, a total of thirteen were statistically significantly associated with Max OXPHOS (Figure 2). Of these, six genes that have preferential localization to the mitochondrion were positively associated with higher Max OXPHOS, including: *SOD2*, *GRX2*, *GRX3*, *PRX3*, *PRX5*, and *TRX2*. Of the genes that have preferential localization to the cytosol and extracellular space, *PRX2* and *PRX6* were positively associated with Max OXPHOS and *GPX3*, *GPX6*, *GRX1*, *PRX1*, and *PRX4* were negatively associated with Max OXPHOS (Supplementary Table 3). Two genes that are exclusively expressed in the testes were not detected via RNAseq and not included in these analyses (*GPX5* and *GPX6*).

**Figure 2.**
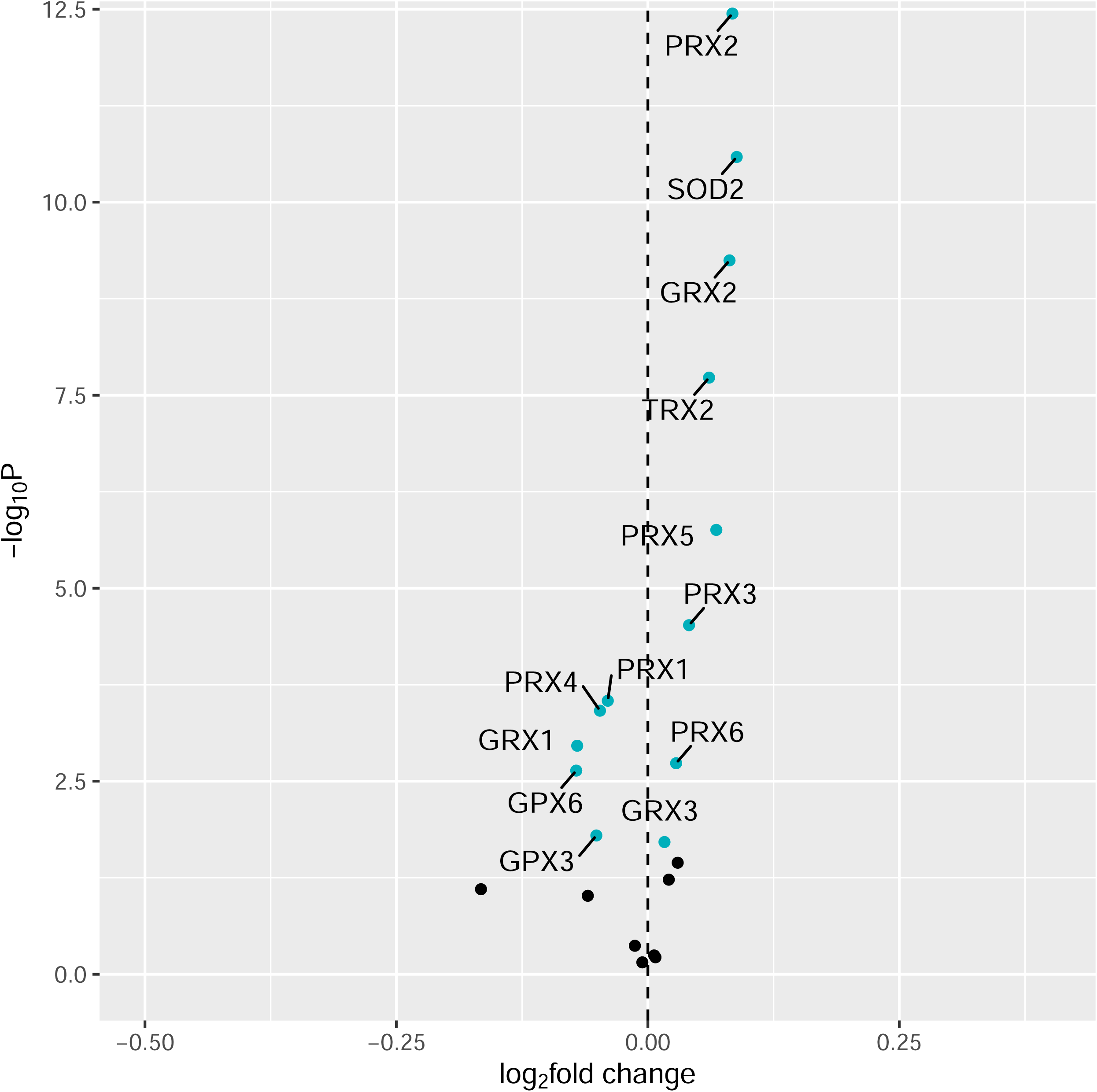
Associations with Max OXPHOS. Volcano plot capturing all statistically significant (p < 0.05 FDR adjusted) genes identified by our models: Each dot represents a gene; the dot color indicates significance level. Base model: gene expression∼Max OXPHOS + age + gender + clinic site + race/ethnicity + height + weight + physical activities + multimorbidity count + sequencing batch.

**Figure 3.**
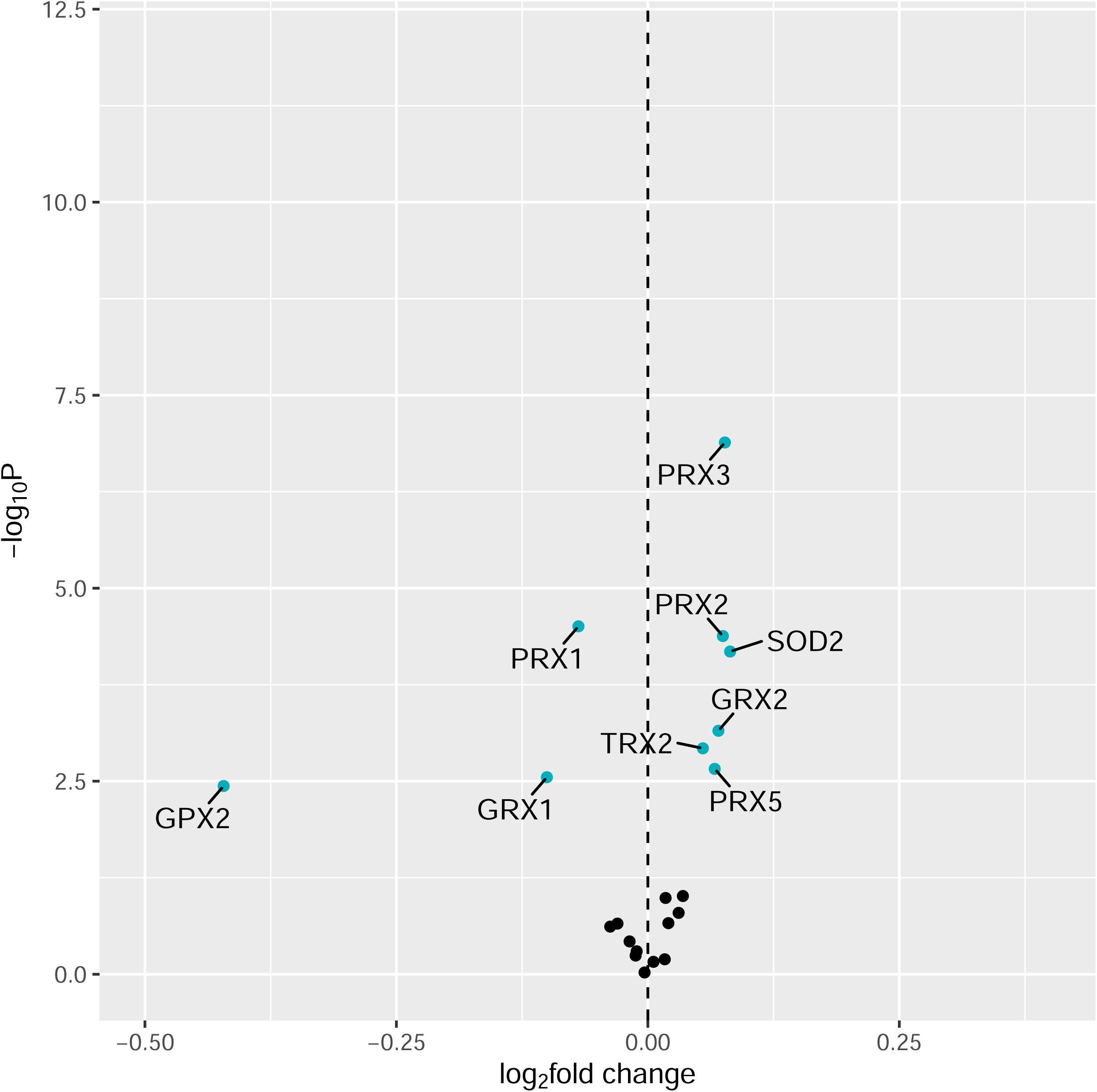
Associations with VO_2_ Peak. Volcano plot capturing all statistically significant (p < 0.05 FDR adjusted) genes identified by our models: Each dot represents a gene; the dot color indicates significance level. Base model: gene expression∼VO_2_ peak + age + gender + clinic site + race/ethnicity + height + weight + physical activities + multimorbidity count + sequencing batch.

### 2.4 Association of VO_2_ peak with gene expression

Most of the oxidative stress genes with preferential localization to the mitochondrion that were associated with Max OXPHOS were also positively associated with higher VO_2_ peak, including: *SOD2*, *GRX2*, *PRX3*, *PRX5*, and *TRX2* (Figure 4). Of the genes with preferential localization to the cytosol and extracellular space, *PRX2* was positively associated with VO_2_ peak and *GPX2*, *GRX1*, and *PRX1* were negatively associated with VO_2_ peak (Supplementary Table 3). A summary of all statistically significant associations with each trait is presented in Figure 4.

**Figure 4.**
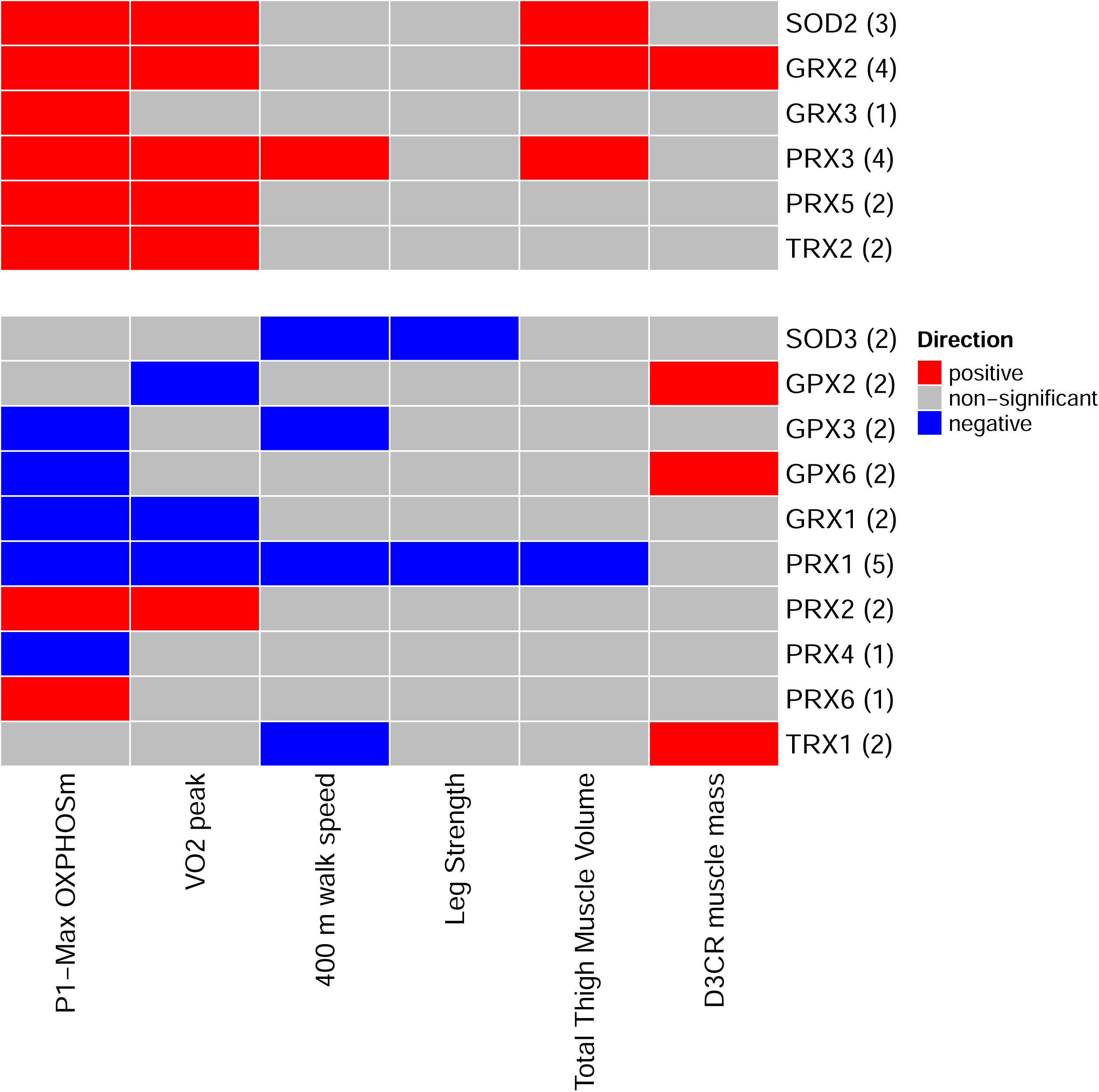
Significant associations with mitochondrial function, physical performance, and muscle mass measures. Heat map capturing all statistically significant (p < 0.05 FDR adjusted) genes identified by our models: each color represents positive (red) or negative (blue) associations.

### 2.5 Association of 400m walk speed with gene expression

A single oxidative stress gene with preferential localization to the mitochondrion, *PRX3*, was positively associated with 400m walking speed. Four genes with preferential localization to the cytosol and extracellular space, *SOD3*, *GPX3*, *PRX1*, and *TRX1* were negatively associated with 400m walking speed (Supplementary Table 5).

### 2.6 Association of leg strength with gene expression

No oxidative stress genes with preferential localization to the mitochondrion were associated with leg strength. Two genes with preferential localization to the cytosol and extracellular space, *SOD3* and *PRX1*, were negatively associated with leg strength (Supplementary Table 6).

### 2.7 Association of total thigh fat free muscle volume with gene expression

Three oxidative stress genes with preferential localization to the mitochondrion, *SOD2*, *GRX2*, and *PRX3*, were positively associated with thigh muscle volume. One gene with preferential localization to the cytosol and extracellular space, *PRX1*, was negatively associated with thigh muscle volume (Supplementary Table 7).

### 2.8 Association of D3CR muscle mass with gene expression

A single oxidative stress gene with preferential localization to the mitochondrion, *GRX2*, and three genes with preferential localization to the cytosol and extracellular space, *GPX2*, *GPX6*, *TRX1*, were positively associated with D3Cr muscle mass (Supplementary Table 8).

## 3. DISCUSSION

In this study of RNA-seq data obtained from muscle biopsies collected from 70–92-year-old individuals, we found that mRNA levels of multiple oxidative stress genes with preferential localization to the mitochondrion were consistently associated with several functional and physical domains. Higher expression of mRNAs that encode *SOD2*, *TRX2*, *PRX3*, *PRX5*, and *GRX2* were positively associated with high levels of mitochondrial respiration in muscle (Max OXPHOS). We originally hypothesized that expression of oxidative stress response genes in the muscle of older persons is not only relevant for mitochondrial function in muscle, but that their expression may be associated with overall fitness, walking speed, strength, and muscle mass. Indeed, five of the six oxidative stress genes with preferential localization to the mitochondrion associated with high muscle mitochondrial muscle function are also positively associated with multiple downstream performance and physical indications, most notably VO_2_ peak (the peak amount of oxygen that an individual utilizes during intense or maximal exercise) which is generally considered the best indicator of cardiovascular fitness and aerobic endurance (Arena et al., 2007) and strongly predicts mortality (Church, Earnest, Skinner, & Blair, 2007). Several of these genes are also positively associated with 400m walking speed and/or measures of muscle mass.

While the mitochondrial and extramitochondrial antioxidant enzymes are well characterized regarding specific functional roles and cellular compartmentalization (Mailloux, 2018; Napolitano et al., 2021) precise molecular mechanisms connecting oxidative stress and the responses to this stress to human mobility have yet to be fully understood. Among the oxidative stress genes with preferential localization to the mitochondrion that were consistently positively associated with multiple traits, several key ‘first responders’ to oxidative stress in the mitochondria were identified. These include *SOD2* (Superoxide Dismutase 2), which is the primary enzyme in the mitochondria responsible for the dismutation of superoxide leading to the formation of hydrogen peroxide (Mailloux, 2018; Napolitano et al., 2021) as well as cytosolic *SOD1* which is also present in the intermembrane space in minor amounts (Okado-Matsumoto & Fridovich, 2001). Active in the mitochondrial matrix, *SOD2* is possibly responsible for broadcasting redox signals generated by mitochondria to distant sites in the cytosol, nucleus, and even outside the cell due to the diffusible nature of hydrogen peroxide (Palma et al., 2020). The remaining associated mitochondrial genes are responsible for metabolizing hydrogen peroxide (*TRX2*, *PRX3*, *PRX5*, and *GRX2*). Among these, *PRX3* may be responsible for metabolizing as much as 90% of the hydrogen peroxide produced by mitochondria (Cox, Winterbourn, & Hampton, 2009). In addition to positive associations with Max OXPHOS and VO_2_ peak, high *PRX3* expression was significantly associated with faster walking speed and higher thigh muscle volume after FDR adjustment and nominally associated with leg strength (p<0.05). Although mitochondria are a significant source of ROS in cells, growing evidence suggests that these organelles may play a less prominent role in oxidant production in contracting skeletal muscles than was previously thought (Powers & Jackson, 2008). Indeed, ROS generation in muscle comes in part from mitochondria but also the NADPH oxidase which generates superoxide intracellularly (Bae, Oh, Rhee, & Yoo, 2011; Martyn, Frederick, von Loehneysen, Dinauer, & Knaus, 2006). Among the genes with preferential localization to the cytosol and extracellular spaces, *PRX1* was negatively associated with numerous measures, including: Max OXPHOS, VO_2_ peak, 400m walking speed, leg strength, and thigh muscle volume. Of the traits examined for associations with cytoplasmic genes, Max OXPHOS yielded the most statistically significant associations but there was inconsistency of direction with some positive and some negative associations.

The remaining genes did not exhibit the consistency of associations across traits like those observed for the mitochondrial oxidative stress response genes. The AREs examined in this study (including superoxide dismutases, glutathione peroxidases, peroxiredoxins, thioredoxins, glutaredoxins) are *Nrf2* target genes (Ma, 2013; Raghunath et al., 2018). The inconsistent directionality of associations we observed between expression of cytoplasm-located genes across traits may in part be explained by impaired *Nrf2* signaling in muscle of older individuals (Zhang et al., 2015). In addition, increased oxidative stress (particularly in mitochondria) can paradoxically increase lifespan in several contexts (Desjardins et al., 2017; Hekimi, 2013; Schaar et al., 2015), suggesting that there is still uncertainty of what constitutes a damaging versus adaptive level of ROS in the cell and that different cellular compartments may respond to elevated ROS in different ways.

Excessive ROS generation can lead to destructive or pathogenic levels of oxidative stress that contribute to muscle dysfunction (Michelucci et al., 2021). Precise regulation of both calcium signaling and ROS production is required for correct skeletal muscle function. However, dysfunctional production of ROS may directly contribute to loss of muscle function, increased damage following muscle rupture, and reduced muscle mass and contractile function in inherited muscle diseases, sarcopenia, and disuse atrophy (Michelucci et al., 2021). Several lines of evidence also connect ROS to muscle atrophy via redox-mediated control of global protein synthesis and proteolysis (Powers, Morton, Ahn, & Smuder, 2016). Disease or inactivity-induced production of ROS in skeletal muscles attenuates protein synthesis by inhibition of *Akt*/*mTORC1* activation resulting in a slower rate of translation (Powers et al., 2016).

Most early studies investigating exercise and free radical production focused on the damaging effects of oxidants in muscle (Powers & Jackson, 2008). However, it has become clear that ROS are involved in modulation of cell signaling pathways and the control of numerous redox-sensitive transcription factors. Furthermore, physiological levels of ROS are essential for optimal force production in skeletal muscle (Powers & Jackson, 2008). Nonetheless, high levels of ROS promote skeletal muscle contractile dysfunction resulting in muscle fatigue (Powers & Jackson, 2008). Exercise only modestly reduces the risk of developing mobility disability (Pahor et al., 2014), requiring the development of new, modifiable intervention targets. For example, Superoxide Dismutase (*SOD*) activity in skeletal muscle is not constant and can be modified by activity patterns. Most studies to date suggest that endurance exercise training promotes 20–112% increases in the activities of both cytosolic *SOD1* and mitochondrial *SOD2* in the exercised muscles (Criswell et al., 1993; Higuchi, Cartier, Chen, & Holloszy, 1985; Lawler, Kwak, Song, & Parker, 2006; Leeuwenburgh, Fiebig, Chandwaney, & Ji, 1994; Leeuwenburgh et al., 1997; Oh-ishi et al., 1997; Powers, Criswell, Lawler, Ji, et al., 1994; Powers, Criswell, Lawler, Martin, et al., 1994; Quintanilha, 1984; Vincent, Powers, Demirel, Coombes, & Naito, 1999; Vincent et al., 2000). Contracting muscles produce oxidants from a variety of cellular locations and maintaining redox homeostasis in muscle fibers requires a network of antioxidant defense mechanisms to reduce the risk of oxidative damage during periods of increased ROS production. As a continuation of the findings presented in this study, future research should validate and leverage these antioxidant mechanisms as modifiable targets for maintaining and improving muscle function and mobility.

This is the first analysis in older adults pairing muscle gene expression and mitochondrial function with several measures of fitness, strength, and muscle mass. Previous studies using muscle biopsies in humans have been small, analyzed only cross-sectional associations between one or two properties and physical performance, and did not include older adults at risk of mobility disability. Moreover, by focusing on well-known oxidative stress response genes we were able to test specific hypotheses regarding the role of oxidative stress genes with preferential localization to the mitochondrion as well as the cytosol and extracellular spaces across mitochondrial function, fitness, mobility, strength and muscle mass. In addition, muscle mass was assessed using the highly accurate D3Cr method. However, the cross-sectional nature of our study precludes evaluation of the directionality of associations. In addition, direct measures of oxidative stress were not assessed. Furthermore, the study participants are from a narrow age range of mostly White race volunteers thus limiting generalizability of findings.

## CONCLUSION

Our results suggest that high antioxidant capacity or low oxidative stress directly may improve mitochondrial function and mobility. Robust and consistent associations across diverse measures of function and muscle mass in older human participants confirms the importance of mitochondrial oxidative stress response as a key determinant of mobility, fitness and function in older men and women. Since higher levels of several mitochondrial oxidative stress response genes were associated with high functioning, they represent promising candidate biomarkers of human mobility in older adults. Identifying specific genes with consistent patterns of association across multiple measures of mitochondrial and physical function are needed to improve our understanding of the biological mechanisms related to human aging and may lead to targeted antioxidant interventions with greater impacts on mobility and independence.

## EXPERIMENTAL PROCEDURES

### 5.1 Study population

The Study of Muscle, Mobility and Aging (SOMMA) is a prospective cohort study of mobility in community-dwelling older adults. Participants for the current study were from the baseline cohort, enrolled between April 2019 and December 2021 (Cummings et al., 2023). SOMMA was conducted at 2 clinical sites: University of Pittsburgh (Pittsburgh, PA) and Wake Forest University School of Medicine (Winston-Salem, NC). Eligible participants were ≥70 years old at enrollment, had a body mass index (BMI) of 18–40 kg/m^2^, and were eligible for magnetic resonance (MR) imaging and a muscle tissue biopsy (Cummings et al., 2023). Individuals were further excluded if they had active cancer or were in the advanced stages of heart failure, renal failure on dialysis, dementia, or Parkinson’s disease. Eligible participants reported ability to walk ¼ mile and climb a flight of stairs and completed a usual pace 400-m walk within 15 minutes during the first day of testing. Participants must have been able to complete the 400 meter walk; those who appeared as they might not be able to complete the 400m walk at the in-person screening visit completed a short distance walk (4 meters) to ensure their walking speed as >=0.6m/s. The study protocol was approved by the Western Institutional Review Board Copernicus Group (WCG IRB; study number 20180764) and all participants provided written informed consent. In brief, baseline testing occurred across 3 separate days of clinic visits that could be up to within 6-8 weeks of each other but had a mean time between visits of 42 days (∼6 weeks). Day 1 included general clinic assessments (eg, physical and cognitive tests; 5 hours), Day 2 included magnetic resonance imaging and Cardiopulmonary Exercise Testing (MR and CPET, 2–3 hours), and Day 3 included fasting specimen and tissue collection (2 hours). There were 879 participants who completed Day 1 of baseline testing and had at least one primary SOMMA measure: CPET, MR imaging, or muscle tissue biopsy.

### 5.2 Demographic, health, and functional measures

#### 5.2.1 Cardiorespiratory fitness (VO_2_ peak)

Cardiorespiratory fitness was measured using gold standard VO_2_ peak (mL/min) from Cardiopulmonary Exercise Testing (CPET). A standardized CPET, using a modified Balke or manual protocol, was administered to participants to measure ventilatory gases, oxygen and carbon dioxide inhaled and exhaled during exercise (Balady et al., 2010). Two slow 5-minute walking tests were conducted before and after the maximal effort test to assess walking energetics at preferred walking speed and a slow fixed speed of 1.5 mph. Participants who were excluded from the maximal effort symptom-limited peak test had acute electrocardiogram (ECG) abnormalities, uncontrolled blood pressure or history of myocardial infarction, unstable angina or angioplasty in the preceding 6 months. Testing for VO_2_ peak began at the participant’s preferred walking speed with incremental rate (0.5 mph) and/or slope (2.5%) increased in 2-minute stages until respiratory exchange ratio, ratio between VCO_2_ and VO_2_, was ≥1.05 and self-reported Borg Rating of Perceived Exertion (Borg, 1982) was ≥17. Blood pressure, pulse oximetry, and ECG were monitored throughout exercise. VO_2_ peak was determined in the BREEZESUITE software (MGC Diagnostics, St. Paul, MN) as the highest 30-second average of VO2 (L/min) achieved. The data were manually reviewed to ensure the correct VO_2_ peak was selected for each participant.

#### 5.2.2 Other measures

Participants are asked to walk at their usual pace for 400 m from which walking speed (m/s) was calculated. Whole-body D_3_Cr muscle mass is measured in participants using a d3-creatine dilution protocol as previously described (Stimpson et al., 2013; Stimpson et al., 2012). Knee extensor leg power was assessed using a Keiser Air 420 exercise machine in the same leg as the muscle biopsy. Resistance to test power was based on determination of the 1 repetition maximum leg extensor strength. Weight was assessed by balance beam or digital scales and height by wall-mounted stadiometers. An approximately 6-minute-long MR scan was taken of the whole body to assess body composition including thigh muscle volume with image processing by AMRA Medical (Linge et al., 2018). The CHAMPS questionnaire (Stewart et al., 2001) was used to assess specific types and the context of physical activities. Participants were asked to self-report physician diagnosis of cancer (excluding nonmelanoma skin cancer), cardiac arrythmia, chronic kidney disease, chronic obstructive pulmonary disease, coronary artery disease, congestive heart failure, depression, diabetes, stroke, and aortic stenosis; from this list a comorbidity count (0, 1, or 2+) was calculated.

#### 5.2.3 Mitochondrial Respiration

Maximal complex I- and II-supported oxidative phosphorylation (Max OXPHOS, also known as State 3 respiration) was measured in permeabilized muscle fiber bundles from biopsies as previously described (Mau et al., 2023).

### 5.3 Gene expression measurement

#### 5.3.1 Skeletal Muscle Biopsy Collection and Processing

Percutaneous biopsies were collected from the mid-thigh *Vastus lateralis* muscle under local anesthesia using a Bergstrom needle with suction (34). Following this, the specimen was blotted dry of blood and interstitial fluid and dissected free of any connective tissue and intermuscular fat. A small amount (∼5-30 mg) was frozen in liquid nitrogen shortly after dissection. Approximately 20 mg of the biopsy specimen was placed into ice-cold BIOPS media (10 mM Ca–EGTA buffer, 0.1 M free calcium, 20 mM imidazole, 20 mM taurine, 50 mM potassium 2-[N-morpholino]-ethanesulfonic acid, 0.5 mM dithiothreitol, 6.56 mM MgCl2, 5.77 mM ATP, and 15 mM phosphocreatine [PCr], pH 7.1) for respirometry, as previously described (22). Myofiber bundles of approximately 2–3 mg were teased apart using a pair of sharp tweezers and a small Petri dish containing ice-cold BIOPS media. After mechanical preparation, myofiber bundles were chemically permeabilized for 30 minutes with saponin (2 mL of 50 μg/mL saponin in ice-cold BIOPS solution) placed on ice and a rocker (25 rpm). Myofiber bundles were washed twice (10 minutes each) with ice-cold MiR05 media (0.5 mM ethylenediaminetetraacetic acid, 3 mM MgCl2·6H2O, 60 mM K-lactobionate, 20 mM taurine, 10 mM KH2PO4, 20 mM N-2-hydroxyethylpiperazine-Nʹ-2-ethanesulfonic acid, 110 mM sucrose, and 1 g/L bovine serum albumin, pH 7.1) on an orbital shaker (25 rpm). The second wash in MiR05 contained blebbistatin (25 μM), a myosin II ATPase inhibitor, that was used to inhibit muscle contraction. Fiber bundle wet weight was determined immediately after permeabilization using an analytical balance (Mettler Toledo, Columbus, OH).

#### 5.3.2 RNA Library Preparation and Sequencing

Total RNA from frozen human skeletal muscle samples (∼5 to 30mg) was prepared using Trizol solution (Invitrogen) according to manufacturer’s direction in 2.0mL Eppendorf safe-lock tubes. Homogenization was performed using the Bullet Blender (NextAdvance, Raymertown NY USA) with an appropriate quantity of stainless-steel beads (autoclaved, 0.5∼2mm, NextAdvance, Raymertown NY USA) at 4°C on Setting 8 (Max is 12) in 30 second bouts. The homogenization step was repeated 5 times for a total of 3 minutes with at least 1 minute break between each bout. The removal of residual genomic DNA was performed by incubating the RNA sample with DNase (AM1907, Thermosci) plus RiboLock RNase inhibitor (EO0381, Thermisci) at 37°C for 25min in a heating block (400rpm). Cleanup of the RNA samples was done using the DNase inactivation reagent following instructions provided by the manufacturer (AM1907, Thermosci). The RNA concentration and integrity were determined by using ThermoSci Nanodrop and Agilent Tapestation.

To prepare RNAseq library, polyA mRNA was isolated from about 250ng total RNA by using NEBNext Poly(A) mRNA magnetic isolation module (E7490L, NEB) and mRNA library constructed by using NEBNext Ultra II directional RNA library Pre Kit for Illumina (E7760L, NEB). Equal molarity of RNAseq libraries were pooled and sequenced on Illumina NovaSeq (2X150bp) to reach 80M reads per sample.

#### 5.3.3 Alignment and Quality Control

The reads from RNA-sequencing were aligned to the Genome Reference Consortium Human Build 38 (GRCh38) using the HISAT2 software (Kim, Paggi, Park, Bennett, & Salzberg, 2019). Duplicated aligned reads were further marked and excluded using the Picardtools software. Expression count data were obtained using the HTseq software (Anders, Pyl, & Huber, 2015). Genes with a total count of ≤ 20 across all samples were filtered out to remove non-expressed genes.

### 5.4 Association analyses

Expression levels of twenty-one oxidative stress response genes including eight with preferential localization to the mitochondrion (*SOD2*, *GPX1*, *GRX2*, *GRX3*, *PRX3*, *PRX5*, *TRX2*, and *CAT*) and thirteen with preferential localization to the cytosol and extracellular spaces (*GPX2*, *GPX3*, *GPX4*, *GPX5*, *GPX6*, *GRX1*, *PRX1*, *PRX2*, *PRX4*, *PRX6*, *SOD1*, *SOD3*, and *TRX1*) were examined. Gene expression associations with traits were identified using negative binomial regression models as implemented by DESeq2 (Love, Huber, & Anders, 2014) in R and adjusted for age, gender, clinic site, race/ethnicity, height, weight, hours per week in all exercise-related activities (CHAMPS), multimorbidity count category, and sequencing batch. DESeq2 uses a negative binomial generalized linear model for differential analysis and applies the Wald test for the significance of GLM coefficients. The Benjamini-Hochberg false discovery rate method was used for P-value adjustment. Genes were considered differentially expressed according to the significance criteria of FDR<0.05. In negative binomial models, traits were modeled using the number of standard deviations (SDs) from each trait’s mean. Consequently, the reported log base 2-fold changes reflect the change in gene expression per one SD unit increase in each trait. Volcano plots were created to visualize the differential expression of RNAs (ENSGs) associated with functional measures. Heat maps were created to summarize significant ENSG associations across all analyzed traits.

## Statement relating to relevant ethics and integrity policies

The study protocol was approved by the Western Institutional Review Board Copernicus Group (WCG IRB; study number 20180764) and all participants provided written informed consent.

## Acknowledgments

For a full list of personnel who contributed to the SOMMA study, please see (Cummings et al., 2023).

## Conflict of Interest statement

S.R.C. is a consultant to Bioage Labs. P.M.Ca. is a consultant to and owns stock in MyoCorps. All other authors declare no conflict of interest.

## Funding statement

SOMMA is funded by the National Institute on Aging (NIA) grant number R01AG059416. Study infrastructure support was funded in part by NIA Claude D. Pepper Older American Independence Centers at University of Pittsburgh (P30AG024827) and Wake Forest University (P30AG021332) and the Clinical and Translational Science Institutes, funded by the National Center for Advancing Translational Science, at Wake Forest University (UL10TR001420).

## Authors’ contributions

Peggy M Cawthon, Paul M Coen, Steven R Cummings, Karyn A Esser, Russell T Hepple, and Gregory J Tranah designed the study. Gregory J Tranah drafted the manuscript. Haley N Barnes, Paul M Coen, Karyn A Esser, Daniel S Evans, and Zhiguang Huo, conducted the genetic analyses. Paul M Coen, Steven R Cummings, Russell T Hepple, Philip A Kramer, and Gregory J Tranah contributed to the interpretation of the results. Haley N Barnes, Peggy M Cawthon, Paul M Coen, Steven R Cummings, Russell T Hepple, Philip A Kramer, and Frederico G S Toledo critically revised the manuscript. All authors reviewed and approved the final version of the manuscript and Haley N Barnes, Zhiguang Huo and Daniel S Evans had full access to the data in the study and accept responsibility to submit for publication.

## Data availability statement

All SOMMA data are publicly available via a web portal. Updated datasets are released approximately every 6 months (https://sommaonline.ucsf.edu/).

## Supplementary information

**Supplemental Table 1:** List of twenty-one oxidative stress response genes, ENSG identifiers, and preferred cellular localizations.

| Gene_symbol | ensembl_gene_id | Cellular localization |
| --- | --- | --- |
| CAT | ENSG00000121691 | Mitochondria |
| GPX1 | ENSG00000233276 | Mitochondria |
| GPX2 | ENSG00000176153 | Extra-mitochondria |
| GPX3 | ENSG00000211445 | Extra-mitochondria |
| GPX4 | ENSG00000167468 | Extra-mitochondria |
| GPX5 | ENSG00000116157 | Extra-mitochondria |
| GPX6 | ENSG00000164294 | Extra-mitochondria |
| GRX1 | ENSG00000173221 | Extra-mitochondria |
| GRX2 | ENSG00000023572 | Mitochondria |
| GRX3 | ENSG00000108010 | Mitochondria |
| PRX1 | ENSG00000117450 | Extra-mitochondria |
| PRX2 | ENSG00000167815 | Extra-mitochondria |
| PRX3 | ENSG00000165672 | Mitochondria |
| PRX4 | ENSG00000123131 | Extra-mitochondria |
| PRX5 | ENSG00000126432 | Mitochondria |
| PRX6 | ENSG00000117592 | Extra-mitochondria |
| SOD1 | ENSG00000142168 | Extra-mitochondria |
| SOD2 | ENSG00000112096 | Mitochondria |
| SOD3 | ENSG00000109610 | Extra-mitochondria |
| TRX1 | ENSG00000136810 | Extra-mitochondria |
| TRX2 | ENSG00000100348 | Mitochondria |

**Supplemental Table 2:** RNAseq quality metrics for each sample, including: number of raw pairs, alignment rate, number of aligned reads, duplication rate, and number of aligned reads deduplicated.

| Sample | Number of raw pairs | Alignment Rate | Number of aligned reads | Duplication Rate | Number of aligned reads deduplicated |
| --- | --- | --- | --- | --- | --- |
| 1 | 96393453 | 95.67% | 184441951 | 78.70% | 39359245 |
| 2 | 100059418 | 96.13% | 192367958 | 68.10% | 61433169 |
| 3 | 93532566 | 96.08% | 179727375 | 68.50% | 56555649 |
| 4 | 90130020 | 95.93% | 172917121 | 62.90% | 64215917 |
| 5 | 93513176 | 95.74% | 179061926 | 66.50% | 60023883 |
| 6 | 93399507 | 96.03% | 179390873 | 78.60% | 38357663 |
| 7 | 91533891 | 96.04% | 175814274 | 75.50% | 43079694 |
| 8 | 95660352 | 95.15% | 182046440 | 86% | 25426123 |
| 9 | 92869196 | 95.60% | 177559973 | 73.80% | 46546318 |
| 10 | 94206508 | 95.61% | 180140177 | 65.40% | 62368154 |
| 11 | 91580510 | 96.24% | 176269851 | 68.30% | 55963786 |
| 12 | 96476311 | 96.31% | 185835269 | 65.40% | 64275793 |
| 13 | 93256753 | 96.06% | 179170646 | 69.90% | 53926180 |
| 14 | 94737694 | 96.51% | 182871302 | 70.80% | 53335997 |
| 15 | 94853873 | 94.61% | 179484404 | 91.60% | 15094298 |
| 16 | 93254346 | 96.10% | 179236781 | 79.80% | 36214863 |
| 17 | 94768451 | 96.06% | 182078220 | 70.30% | 53994168 |
| 18 | 92703785 | 96.36% | 178650402 | 61.20% | 69327789 |
| 19 | 92792742 | 96.33% | 178781101 | 65.80% | 61190811 |
| 20 | 95448973 | 96.50% | 184214920 | 71.60% | 52242426 |
| 21 | 96386549 | 95.70% | 184490763 | 65.20% | 64208390 |
| 22 | 94049712 | 95.77% | 180144045 | 85.40% | 26374250 |
| 23 | 92534259 | 95.52% | 176781852 | 82% | 31751068 |
| 24 | 96136369 | 95.47% | 183567705 | 77.70% | 40894545 |
| 25 | 92684795 | 96.66% | 179169760 | 79% | 37553138 |
| 26 | 94274385 | 95.62% | 180291886 | 81% | 34329087 |
| 27 | 95238852 | 95.32% | 181565967 | 81.80% | 32954237 |
| 28 | 94658476 | 95.26% | 180343006 | 89.90% | 18235077 |
| 29 | 92917699 | 95.43% | 177341522 | 75.20% | 43975639 |
| 30 | 97826542 | 96.21% | 188243115 | 72.50% | 51785757 |
| 31 | 93211235 | 95.45% | 177942884 | 83.40% | 29533846 |
| 32 | 95606463 | 95.69% | 182973610 | 75.20% | 45433035 |
| 33 | 95441961 | 96.31% | 183849801 | 74.60% | 46701458 |
| 34 | 96075319 | 96.12% | 184697195 | 74.20% | 47673981 |
| 35 | 95074060 | 96.35% | 183208134 | 70.80% | 53417857 |
| 36 | 91693188 | 95.82% | 175727627 | 77.60% | 39393394 |
| 37 | 92617017 | 92.67% | 171647448 | 90.10% | 16974404 |
| 38 | 95760744 | 95.74% | 183363658 | 78% | 40363306 |
| 39 | 93959617 | 94.95% | 178419945 | 82.10% | 32008520 |
| 40 | 97687230 | 95.69% | 186945742 | 72.80% | 50891281 |
| 41 | 95290985 | 96.08% | 183118608 | 76.10% | 43755133 |
| 42 | 96405400 | 96.10% | 185288591 | 71.30% | 53153408 |
| 43 | 91618248 | 95.70% | 175361499 | 74% | 45638953 |
| 44 | 90604635 | 95.59% | 173221766 | 79.70% | 35235993 |
| 45 | 91908525 | 94.24% | 173221121 | 73.90% | 45147052 |
| 46 | 93339668 | 95.45% | 178183219 | 86.60% | 23847280 |
| 47 | 93228195 | 95.38% | 177838787 | 80.10% | 35379447 |
| 48 | 94185936 | 95.70% | 180277596 | 85.90% | 25478974 |
| 49 | 93076777 | 95.60% | 177965955 | 74.90% | 44623024 |
| 50 | 92853231 | 95.51% | 177372625 | 67.50% | 57638727 |
| 51 | 95924802 | 95.83% | 183844971 | 83.20% | 30890475 |
| 52 | 91701902 | 93.61% | 171676642 | 88.50% | 19794352 |
| 53 | 93978044 | 95.60% | 179680494 | 82.20% | 31952553 |
| 54 | 93498088 | 94.84% | 177339169 | 85.90% | 25052790 |
| 55 | 91373701 | 95.10% | 173792287 | 86% | 24339052 |
| 56 | 92633161 | 94.21% | 174537705 | 92.40% | 13282764 |
| 57 | 93632029 | 94.82% | 177555337 | 74.70% | 44895907 |
| 58 | 96935810 | 96.11% | 186322556 | 66.40% | 62655363 |
| 59 | 95970326 | 95.97% | 184214134 | 68.70% | 57608266 |
| 60 | 92800167 | 95.46% | 177179393 | 86.60% | 23666468 |
| 61 | 102699034 | 96.30% | 197801041 | 64.90% | 69504546 |
| 62 | 92654946 | 94.91% | 175870882 | 85% | 26296527 |
| 63 | 92637261 | 94.25% | 174620099 | 85.40% | 25547736 |
| 64 | 93933534 | 94.91% | 178301144 | 86% | 24893302 |
| 65 | 94850969 | 94.69% | 179622664 | 91.10% | 15967433 |
| 66 | 93576231 | 95.41% | 178562087 | 80.90% | 34065450 |
| 67 | 90485671 | 95.26% | 172384541 | 75.90% | 41587116 |
| 68 | 93379297 | 94.19% | 175909909 | 91.20% | 15397217 |
| 69 | 96498988 | 95.91% | 185101771 | 73.10% | 49753846 |
| 70 | 71461019 | 95.40% | 136344256 | 59.30% | 55504737 |
| 71 | 93752448 | 95.60% | 179248936 | 59.20% | 73073568 |
| 72 | 86411126 | 96.23% | 166306753 | 58.30% | 69376925 |
| 73 | 83235259 | 96.26% | 160250568 | 55.50% | 71262583 |
| 74 | 81261596 | 96.36% | 156615091 | 60.10% | 62471112 |
| 75 | 97783727 | 95.85% | 187450816 | 57.10% | 80348879 |
| 76 | 83767720 | 95.75% | 160407526 | 57.60% | 68092398 |
| 77 | 86729806 | 95.31% | 165318863 | 57.60% | 70036456 |
| 78 | 110942864 | 95.51% | 211919823 | 57% | 91162871 |
| 79 | 104966827 | 96.19% | 201945250 | 56% | 88772846 |
| 80 | 98253075 | 96.48% | 189598150 | 65.60% | 65198970 |
| 81 | 116927639 | 96.27% | 225129926 | 56.50% | 97959452 |
| 82 | 112176949 | 96.19% | 215797160 | 58.40% | 89813138 |
| 83 | 98534039 | 95.80% | 188796585 | 53.50% | 87847469 |
| 84 | 108954720 | 95.86% | 208892196 | 53.40% | 97416327 |
| 85 | 84097255 | 95.78% | 161090321 | 57.10% | 69129756 |
| 86 | 95236027 | 95.90% | 182663085 | 61.60% | 70206810 |
| 87 | 88597819 | 95.87% | 169884224 | 55.30% | 75955704 |
| 88 | 96328455 | 96.12% | 185180033 | 57.80% | 78177200 |
| 89 | 90461109 | 96.09% | 173855662 | 56.80% | 75088574 |
| 90 | 102870253 | 95.60% | 196695477 | 57.10% | 84377364 |
| 91 | 108655599 | 96.07% | 208762677 | 55% | 93925156 |
| 92 | 106334623 | 95.39% | 202873100 | 58% | 85169516 |
| 93 | 95273543 | 96.52% | 183917484 | 59.50% | 74534598 |
| 94 | 92910303 | 95.10% | 176722146 | 61.70% | 67734224 |
| 95 | 85877520 | 95.74% | 164431755 | 57.30% | 70181106 |
| 96 | 101420521 | 96.01% | 194746668 | 57.80% | 82270980 |
| 97 | 106560498 | 95.79% | 204151071 | 60.10% | 81465013 |
| 98 | 88052217 | 95.85% | 168790927 | 61.40% | 65182021 |
| 99 | 86639281 | 95.98% | 166305148 | 60.20% | 66160365 |
| 100 | 110166267 | 96.12% | 211781602 | 58.40% | 88101366 |
| 101 | 68517291 | 95.90% | 131415364 | 58.40% | 54667509 |
| 102 | 82185872 | 89.49% | 147099825 | 56.90% | 63463056 |
| 103 | 81881951 | 91.49% | 149826924 | 51.60% | 72577249 |
| 104 | 79982310 | 92.80% | 148454721 | 54.20% | 68030223 |
| 105 | 80404905 | 93.43% | 150237431 | 54.10% | 68971717 |
| 106 | 80821283 | 92.89% | 150151720 | 56.80% | 64805299 |
| 107 | 81203942 | 92.14% | 149636180 | 52.50% | 71064306 |
| 108 | 83124572 | 93.65% | 155692925 | 55% | 70035174 |
| 109 | 83732758 | 91.81% | 153750133 | 56% | 67697013 |
| 110 | 85425657 | 93.21% | 159252490 | 55.20% | 71368045 |
| 111 | 83608132 | 92.59% | 154821249 | 55.10% | 69477695 |
| 112 | 80057039 | 92.61% | 148280864 | 51.40% | 72014599 |
| 113 | 80421460 | 92.78% | 149231205 | 55.70% | 66145867 |
| 114 | 79661371 | 93.39% | 148792790 | 52.40% | 70837393 |
| 115 | 78552545 | 93.67% | 147161472 | 49.60% | 74116342 |
| 116 | 80742166 | 93.70% | 151310619 | 53.80% | 69932072 |
| 117 | 81952954 | 93.45% | 153170108 | 51.40% | 74467136 |
| 118 | 84605721 | 92.48% | 156488222 | 53.50% | 72754366 |
| 119 | 83317876 | 93.43% | 155693857 | 54.60% | 70715421 |
| 120 | 84072070 | 92.31% | 155213840 | 55.10% | 69658730 |
| 121 | 78879445 | 93.85% | 148060431 | 54.30% | 67613149 |
| 122 | 81065245 | 93.13% | 150987465 | 52.90% | 71065006 |
| 123 | 80217494 | 93.74% | 150386891 | 52.60% | 71283062 |
| 124 | 78825058 | 93.19% | 146907604 | 53% | 69007099 |
| 125 | 82173370 | 92.22% | 151561681 | 56% | 66722801 |
| 126 | 84325245 | 91.89% | 154967259 | 55.80% | 68529272 |
| 127 | 83005575 | 92.12% | 152925352 | 52.40% | 72862136 |
| 128 | 79764300 | 92.28% | 147216468 | 54.60% | 66881432 |
| 129 | 80033284 | 93.50% | 149658311 | 53.60% | 69425984 |
| 130 | 83094845 | 92.07% | 153013625 | 56% | 67339202 |
| 131 | 81833515 | 93.24% | 152604746 | 53.20% | 71429012 |
| 132 | 81767329 | 92.75% | 151672136 | 54.50% | 69022261 |
| 133 | 86613650 | 93.83% | 162536626 | 50.90% | 79868855 |
| 134 | 84575964 | 93.04% | 157374691 | 52.80% | 74277949 |
| 135 | 84551330 | 93.20% | 157608706 | 57.30% | 67364855 |
| 136 | 79377942 | 92.20% | 146379254 | 54.60% | 66496956 |
| 137 | 82220781 | 93.55% | 153841128 | 53.80% | 71087389 |
| 138 | 84490367 | 93.66% | 158262891 | 49% | 80737589 |
| 139 | 79135730 | 93.46% | 147925972 | 54.60% | 67200337 |
| 140 | 79315800 | 93.29% | 147985482 | 53.80% | 68321650 |
| 141 | 83183251 | 93.78% | 156011569 | 56.20% | 68318618 |
| 142 | 80033861 | 93.20% | 149184350 | 54.50% | 67886689 |
| 143 | 81365287 | 93.54% | 152211890 | 53.10% | 71334270 |
| 144 | 78302039 | 93.65% | 146653385 | 52% | 70451521 |
| 145 | 78250641 | 93.72% | 146677528 | 50.20% | 72978169 |
| 146 | 88982141 | 93.56% | 166509772 | 51.30% | 81144529 |
| 147 | 83465583 | 93.76% | 156512508 | 52.70% | 73991949 |
| 148 | 78062842 | 93.16% | 145453918 | 54.90% | 65605230 |
| 149 | 82988051 | 93.61% | 155366184 | 54.10% | 71328165 |
| 150 | 81862895 | 93.18% | 152566757 | 53.50% | 70874912 |
| 151 | 83357795 | 92.99% | 155033190 | 53.90% | 71547631 |
| 152 | 78541474 | 92.61% | 145477010 | 55.20% | 65238613 |
| 153 | 79658492 | 93.21% | 148502581 | 54% | 68314118 |
| 154 | 83568623 | 93.12% | 155630779 | 51.10% | 76070555 |
| 155 | 82216893 | 93.36% | 153507981 | 54.80% | 69417332 |
| 156 | 80284841 | 93.01% | 149342427 | 53.80% | 69032431 |
| 157 | 81855521 | 92.63% | 151649262 | 57.70% | 64177810 |
| 158 | 83777511 | 92.15% | 154404604 | 56.40% | 67384639 |
| 159 | 76397124 | 92.84% | 141852133 | 52.10% | 67974888 |
| 160 | 76699266 | 93.64% | 143635884 | 52.50% | 68190049 |
| 161 | 81119697 | 92.98% | 150848135 | 57.30% | 64381281 |
| 162 | 80828343 | 93.88% | 151762556 | 53.70% | 70201572 |
| 163 | 81754170 | 92.54% | 151315363 | 54.40% | 68934509 |
| 164 | 77182270 | 92.88% | 143378696 | 54.50% | 65188366 |
| 165 | 83277951 | 91.93% | 153115251 | 54.80% | 69242593 |
| 166 | 81434647 | 93.24% | 151858802 | 53% | 71313059 |
| 167 | 80097377 | 93.07% | 149095520 | 56% | 65587909 |
| 168 | 80351222 | 92.75% | 149049068 | 55.40% | 66477867 |
| 169 | 81065470 | 92.60% | 150136734 | 56.50% | 65376792 |
| 170 | 79732292 | 92.77% | 147932802 | 51.20% | 72190979 |
| 171 | 81364923 | 92.82% | 151041094 | 54.70% | 68468523 |
| 172 | 84217555 | 92.71% | 156156243 | 53.40% | 72719070 |
| 173 | 82659700 | 91.86% | 151855951 | 58.90% | 62427158 |
| 174 | 77764144 | 92.67% | 144122794 | 54.70% | 65326768 |
| 175 | 78458276 | 93.61% | 146894807 | 53.40% | 68434532 |
| 176 | 82281245 | 92.59% | 152375583 | 54.70% | 69096970 |
| 177 | 81723750 | 93.04% | 152071478 | 53.70% | 70467804 |
| 178 | 92966002 | 93.08% | 173061954 | 53% | 81409313 |
| 179 | 108642167 | 92.90% | 201857878 | 54.50% | 91931884 |
| 180 | 110042963 | 93.11% | 204915230 | 55.40% | 91326091 |
| 181 | 80782419 | 91.74% | 148214005 | 55.40% | 66133720 |
| 182 | 84525342 | 93.26% | 157656520 | 54.20% | 72184934 |
| 183 | 83707351 | 92.93% | 155580510 | 56.70% | 67374897 |
| 184 | 81396801 | 92.67% | 150863483 | 54.20% | 69072120 |
| 185 | 79461855 | 92.93% | 147683370 | 53.60% | 68516893 |
| 186 | 82835786 | 93.45% | 154824888 | 56.10% | 68011860 |
| 187 | 80121046 | 91.52% | 146653254 | 56.20% | 64198018 |
| 188 | 80677720 | 93.08% | 150191167 | 51.70% | 72467988 |
| 189 | 73976840 | 92.95% | 137522860 | 55.50% | 61139754 |
| 190 | 84296590 | 92.68% | 156244470 | 54.30% | 71397717 |
| 191 | 79357053 | 92.05% | 146094379 | 57.40% | 62260719 |
| 192 | 83738743 | 92.78% | 155381196 | 55.10% | 69733453 |
| 193 | 81718359 | 93.00% | 151991385 | 55.70% | 67276304 |
| 194 | 82041039 | 92.73% | 152145911 | 53.70% | 70411580 |
| 195 | 80029302 | 93.27% | 149286808 | 54.60% | 67736131 |
| 196 | 80785074 | 92.28% | 149102807 | 58.20% | 62289713 |
| 197 | 83843652 | 89.95% | 150834458 | 55.60% | 66959638 |
| 198 | 83590981 | 90.54% | 151374766 | 54.40% | 69016209 |
| 199 | 83437723 | 90.56% | 151119766 | 61.30% | 58460828 |
| 200 | 83200552 | 90.82% | 151130394 | 58.30% | 63046406 |
| 201 | 85626471 | 91.26% | 156284569 | 54.70% | 70851462 |
| 202 | 85639782 | 91.15% | 156127532 | 55.30% | 69759580 |
| 203 | 82537377 | 92.16% | 152128868 | 54.80% | 68808968 |
| 204 | 85570850 | 91.07% | 155854052 | 55.50% | 69431392 |
| 205 | 85934669 | 88.37% | 151874286 | 57.70% | 64291224 |
| 206 | 82777122 | 89.44% | 148070369 | 56.30% | 64655527 |
| 207 | 84965709 | 91.77% | 155946997 | 60.90% | 60917009 |
| 208 | 83933360 | 91.66% | 153860969 | 56.30% | 67229061 |
| 209 | 84894553 | 91.66% | 155625165 | 54.60% | 70729417 |
| 210 | 86259859 | 91.23% | 157396509 | 57.40% | 67053274 |
| 211 | 84346300 | 90.21% | 152170674 | 60.30% | 60352643 |
| 212 | 81677653 | 92.44% | 151003784 | 54% | 69511530 |
| 213 | 84048781 | 90.99% | 152954587 | 54.50% | 69606002 |
| 214 | 83654135 | 91.72% | 153456971 | 56.10% | 67299283 |
| 215 | 82130420 | 91.59% | 150453661 | 55.80% | 66435733 |
| 216 | 86286063 | 92.62% | 159833996 | 55.20% | 71548012 |
| 217 | 81626143 | 92.37% | 150800166 | 53.50% | 70150436 |
| 218 | 88346127 | 92.33% | 163143773 | 61.10% | 63421969 |
| 219 | 84865918 | 91.48% | 155278413 | 59.90% | 62191416 |
| 220 | 84973197 | 91.01% | 154664003 | 57.10% | 66420352 |
| 221 | 85254795 | 91.64% | 156259490 | 52.70% | 73942216 |
| 222 | 83789654 | 91.01% | 152511134 | 58.80% | 62864169 |
| 223 | 95815599 | 92.47% | 177201844 | 57.80% | 74694621 |
| 224 | 81589169 | 92.29% | 150602813 | 53.40% | 70253955 |
| 225 | 84008407 | 93.45% | 157012490 | 54.40% | 71645670 |
| 226 | 91458190 | 92.84% | 169827777 | 52% | 81521417 |
| 227 | 85816384 | 92.18% | 158208155 | 51.90% | 76103627 |
| 228 | 85124387 | 92.51% | 157502606 | 53.30% | 73491302 |
| 229 | 95718199 | 92.66% | 177393846 | 50.90% | 87012401 |
| 230 | 88765558 | 91.84% | 163040208 | 50.20% | 81131345 |
| 231 | 85307518 | 92.10% | 157139563 | 47.20% | 82950928 |
| 232 | 85215525 | 92.17% | 157078878 | 52.20% | 75159281 |
| 233 | 92867670 | 92.78% | 172320565 | 50.30% | 85711818 |
| 234 | 95802997 | 92.10% | 176464214 | 51% | 86392608 |
| 235 | 90056750 | 92.42% | 166462816 | 52.40% | 79208945 |
| 236 | 88674254 | 92.26% | 163617746 | 49.10% | 83319295 |
| 237 | 87963713 | 93.34% | 164213747 | 49.40% | 83028930 |
| 238 | 92809866 | 92.70% | 172061848 | 51.70% | 83186393 |
| 239 | 87668758 | 92.36% | 161935944 | 54.80% | 73257571 |
| 240 | 83691132 | 93.13% | 155876722 | 52.30% | 74285916 |
| 241 | 87620635 | 93.43% | 163722214 | 48.50% | 84303328 |
| 242 | 87370476 | 93.83% | 163955893 | 51.50% | 79457633 |
| 243 | 85183296 | 93.51% | 159304394 | 48.90% | 81388632 |
| 244 | 85305143 | 91.00% | 155259108 | 59.40% | 62964001 |
| 245 | 87686269 | 91.76% | 160913835 | 50.10% | 80229213 |
| 246 | 85759147 | 92.88% | 159301455 | 45.80% | 86380616 |
| 247 | 86761827 | 92.46% | 160444212 | 52.30% | 76597750 |
| 248 | 95574789 | 93.14% | 178043997 | 53.60% | 82642295 |
| 249 | 84799240 | 91.03% | 154390548 | 58.90% | 63417450 |
| 250 | 92994203 | 92.25% | 171580708 | 54.90% | 77303002 |
| 251 | 90002275 | 93.51% | 168319214 | 52.50% | 79979804 |
| 252 | 93496332 | 92.41% | 172791290 | 53.20% | 80942510 |
| 253 | 93429096 | 93.13% | 174026090 | 53.40% | 81155282 |
| 254 | 92210444 | 93.87% | 173118774 | 49.60% | 87229532 |
| 255 | 86830309 | 93.01% | 161524479 | 51.20% | 78790992 |
| 256 | 85668489 | 93.69% | 160529297 | 53.20% | 75101647 |
| 257 | 82811822 | 93.50% | 154851807 | 56.70% | 67098167 |
| 258 | 92210607 | 92.75% | 171043151 | 56.20% | 74920206 |
| 259 | 86670973 | 92.98% | 161175596 | 54% | 74204742 |
| 260 | 85317863 | 92.59% | 157985436 | 53.60% | 73349237 |
| 261 | 84376356 | 93.29% | 157430753 | 52% | 75551090 |
| 262 | 88846406 | 93.69% | 166474759 | 52.80% | 78535808 |
| 263 | 84250864 | 93.37% | 157323970 | 49.90% | 78810698 |
| 264 | 83884503 | 93.41% | 156707758 | 55.70% | 69362822 |
| 265 | 84039086 | 93.25% | 156727952 | 58.10% | 65613249 |
| 266 | 85755986 | 93.72% | 160745223 | 52.40% | 76444502 |
| 267 | 84323917 | 93.02% | 156873725 | 55.10% | 70465299 |
| 268 | 86565553 | 93.53% | 161926567 | 50.80% | 79604802 |
| 269 | 86567025 | 93.16% | 161289477 | 50.10% | 80523325 |
| 270 | 88494339 | 92.87% | 164361568 | 52% | 78929966 |
| 271 | 86623009 | 93.02% | 161151152 | 51.20% | 78600588 |
| 272 | 86327648 | 93.10% | 160745980 | 54% | 73918147 |
| 273 | 83141346 | 93.24% | 155036034 | 53.30% | 72386235 |
| 274 | 83302444 | 93.24% | 155343331 | 60.30% | 61680278 |
| 275 | 84909067 | 93.47% | 158736296 | 53.10% | 74517144 |
| 276 | 94998934 | 93.04% | 176782669 | 54.20% | 80973713 |
| 277 | 85473106 | 91.72% | 156783969 | 48% | 81529587 |
| 278 | 84419283 | 93.08% | 157151408 | 49.60% | 79147797 |
| 279 | 84339155 | 92.54% | 156093345 | 54.10% | 71659275 |
| 280 | 82898656 | 93.49% | 155005772 | 57.10% | 66469529 |
| 281 | 88618338 | 92.36% | 163687680 | 55.50% | 72864712 |
| 282 | 84028068 | 92.83% | 155998753 | 55.40% | 69596791 |
| 283 | 86880065 | 92.54% | 160795831 | 54.20% | 73623117 |
| 284 | 84845899 | 93.53% | 158711551 | 51.20% | 77485314 |
| 285 | 84860629 | 92.11% | 156330197 | 52.40% | 74379075 |
| 286 | 84257108 | 93.42% | 157421881 | 52.40% | 74920353 |
| 287 | 84280613 | 93.66% | 157878588 | 50.10% | 78725756 |
| 288 | 84861735 | 93.52% | 158721939 | 53.70% | 73496627 |
| 289 | 85386584 | 93.20% | 159158664 | 51.50% | 77183783 |
| 290 | 85547278 | 91.97% | 157357004 | 64.40% | 56086510 |
| 291 | 85506856 | 92.17% | 157623630 | 50.60% | 77883014 |
| 292 | 87465435 | 92.87% | 162465151 | 48.70% | 83288893 |
| 293 | 89476865 | 93.44% | 167220449 | 50.30% | 83141110 |
| 294 | 82110594 | 93.53% | 153591414 | 55% | 69078281 |
| 295 | 84093778 | 92.59% | 155729679 | 60.40% | 61678538 |
| 296 | 83423489 | 93.01% | 155185118 | 59.60% | 62749001 |
| 297 | 85932540 | 93.29% | 160337553 | 59.40% | 65131305 |
| 298 | 83538416 | 93.34% | 155953809 | 52.40% | 74289150 |
| 299 | 84091188 | 93.52% | 157277665 | 49.20% | 79941873 |
| 300 | 90069946 | 93.01% | 167540972 | 52.10% | 80254592 |
| 301 | 84061111 | 93.66% | 157457891 | 50.80% | 77394403 |
| 302 | 83155272 | 93.16% | 154932286 | 57.50% | 65789921 |
| 303 | 83585064 | 92.55% | 154711276 | 56.30% | 67639225 |
| 304 | 84628574 | 93.46% | 158188844 | 55.90% | 69742868 |
| 305 | 83011782 | 92.14% | 152977493 | 54.20% | 70059215 |
| 306 | 81280441 | 93.78% | 152451484 | 50.80% | 74998130 |
| 307 | 84245261 | 92.76% | 156299058 | 54% | 71892451 |
| 308 | 84725802 | 92.59% | 156893654 | 51.20% | 76591677 |
| 309 | 84866329 | 92.66% | 157282114 | 52.60% | 74575212 |
| 310 | 85688224 | 92.84% | 159099875 | 53.30% | 74358069 |
| 311 | 85437828 | 91.87% | 156986962 | 62.70% | 58478149 |
| 312 | 84433153 | 92.86% | 156816220 | 55.70% | 69523283 |
| 313 | 84621049 | 92.92% | 157251935 | 53.20% | 73551058 |
| 314 | 83144014 | 93.91% | 156162315 | 50.30% | 77646238 |
| 315 | 83362769 | 92.86% | 154822328 | 50.70% | 76272766 |
| 316 | 84481479 | 93.31% | 157655895 | 49.30% | 79968258 |
| 317 | 86230336 | 93.73% | 161655502 | 50.80% | 79536622 |
| 318 | 84799431 | 92.73% | 157261019 | 49.80% | 78957526 |
| 319 | 78405835 | 93.18% | 146109354 | 48% | 75928263 |
| 320 | 88892787 | 93.76% | 166692480 | 50.80% | 82020430 |
| 321 | 85070862 | 93.32% | 158769321 | 48% | 82521937 |
| 322 | 84947408 | 94.30% | 160214849 | 50.80% | 78829492 |
| 323 | 92407695 | 93.73% | 173228551 | 52.50% | 82368708 |
| 324 | 84891874 | 94.27% | 160048590 | 50.70% | 78926705 |
| 325 | 85709910 | 94.39% | 161804370 | 51.50% | 78417315 |
| 326 | 83101247 | 94.01% | 156241366 | 48.50% | 80410232 |
| 327 | 82308793 | 93.72% | 154274395 | 52.10% | 73843575 |
| 328 | 87109225 | 94.00% | 163759092 | 51% | 80301894 |
| 329 | 86532923 | 92.20% | 159572561 | 50.90% | 78415267 |
| 330 | 90737882 | 90.54% | 164315402 | 54.90% | 74053890 |
| 331 | 83355319 | 92.29% | 153862208 | 50.30% | 76523033 |
| 332 | 85026830 | 93.60% | 159169119 | 51.60% | 76985647 |
| 333 | 83133743 | 92.89% | 154448461 | 52.50% | 73314153 |
| 334 | 81974796 | 92.52% | 151679531 | 49.70% | 76328624 |
| 335 | 85411349 | 93.16% | 159131845 | 48.70% | 81630732 |
| 336 | 87230169 | 89.93% | 156896035 | 51.20% | 76597809 |
| 337 | 86522843 | 93.28% | 161421146 | 52.10% | 77300193 |
| 338 | 83912636 | 92.91% | 155918473 | 47.40% | 81939343 |
| 339 | 96064117 | 93.21% | 179087734 | 55.50% | 79780353 |
| 340 | 91874480 | 92.19% | 169398960 | 54.10% | 77734137 |
| 341 | 90458184 | 93.45% | 169069924 | 54.20% | 77368105 |
| 342 | 92262874 | 92.32% | 170355177 | 56.30% | 74361329 |
| 343 | 88623713 | 93.33% | 165428691 | 54.40% | 75387321 |
| 344 | 87207805 | 91.69% | 159918728 | 62.10% | 60610291 |
| 345 | 91004676 | 91.07% | 165761485 | 65.50% | 57159741 |
| 346 | 90916275 | 93.80% | 170560323 | 54.10% | 78288436 |
| 347 | 82776921 | 91.55% | 151571920 | 65.70% | 52028736 |
| 348 | 92768155 | 91.99% | 170681073 | 52.80% | 80587319 |
| 349 | 87856604 | 91.93% | 161536091 | 56% | 71121274 |
| 350 | 94246257 | 93.71% | 176642932 | 54% | 81343271 |
| 351 | 90609020 | 92.28% | 167228668 | 60.30% | 66404884 |
| 352 | 96498298 | 92.76% | 179016244 | 60% | 71599729 |
| 353 | 90315456 | 92.62% | 167293252 | 57.70% | 70834030 |
| 354 | 91413142 | 91.00% | 166373604 | 59.60% | 67260982 |
| 355 | 87915585 | 92.42% | 162503027 | 61.70% | 62305118 |
| 356 | 91125683 | 92.91% | 169336075 | 57.40% | 72191861 |
| 357 | 91398473 | 92.76% | 169571520 | 54.20% | 77661393 |
| 358 | 95481994 | 91.81% | 175329119 | 60.40% | 69382289 |
| 359 | 110818214 | 92.60% | 205241457 | 57.60% | 87117111 |
| 360 | 94806068 | 92.95% | 176246636 | 54.70% | 79789029 |
| 361 | 89836893 | 92.71% | 166580194 | 56.10% | 73142972 |
| 362 | 98638711 | 93.23% | 183921845 | 56.70% | 79659069 |
| 363 | 93853377 | 92.12% | 172917042 | 55.50% | 77019448 |
| 364 | 100275484 | 91.79% | 184078267 | 58.40% | 76604986 |
| 365 | 92495429 | 93.01% | 172062371 | 54.80% | 77718704 |
| 366 | 88715899 | 92.84% | 164731494 | 55.50% | 73260860 |
| 367 | 92461693 | 93.31% | 172556715 | 55.80% | 76329898 |
| 368 | 102100512 | 92.74% | 189382528 | 54% | 87061978 |
| 369 | 90504044 | 92.70% | 167798855 | 60.10% | 66899554 |
| 370 | 89709289 | 93.94% | 168552896 | 52.20% | 80537509 |
| 371 | 87126691 | 93.41% | 162764208 | 60.10% | 64993578 |
| 372 | 84451594 | 93.81% | 158447618 | 54.30% | 72379951 |
| 373 | 92484999 | 93.30% | 172582788 | 56.20% | 75514636 |
| 374 | 95069991 | 92.37% | 175627025 | 53.60% | 81522149 |
| 375 | 91417795 | 93.66% | 171251489 | 54.30% | 78191020 |
| 376 | 87960991 | 90.90% | 159916106 | 59.70% | 64483465 |
| 377 | 99886475 | 93.44% | 186660828 | 57.40% | 79514036 |
| 378 | 87890978 | 92.47% | 162553511 | 57.30% | 69367802 |
| 379 | 92017579 | 93.07% | 171275765 | 61.20% | 66437963 |
| 380 | 87881414 | 93.31% | 163996843 | 54.10% | 75296785 |
| 381 | 88237604 | 93.83% | 165581376 | 52.10% | 79392676 |
| 382 | 87808127 | 93.01% | 163340130 | 55.70% | 72401527 |
| 383 | 91892021 | 94.02% | 172792672 | 52.20% | 82574586 |
| 384 | 86824993 | 93.68% | 162683119 | 53.70% | 75385841 |
| 385 | 83169708 | 92.44% | 153768393 | 64.50% | 54605416 |
| 386 | 92957654 | 91.89% | 170829216 | 61.60% | 65562022 |
| 387 | 90627760 | 93.92% | 170231373 | 63.60% | 61998824 |
| 388 | 94159915 | 93.71% | 176478389 | 53.60% | 81955000 |
| 389 | 93895734 | 92.31% | 173357587 | 67% | 57291911 |
| 390 | 93110943 | 91.98% | 171288798 | 65.50% | 59067248 |
| 391 | 91179562 | 93.39% | 170307295 | 60.70% | 66994666 |
| 392 | 94007538 | 94.13% | 176984606 | 57% | 76037137 |
| 393 | 92783466 | 91.90% | 170532129 | 65.30% | 59175784 |
| 394 | 93347423 | 93.30% | 174193583 | 54.10% | 80031876 |
| 395 | 93719900 | 92.07% | 172581611 | 56.80% | 74499816 |
| 396 | 94289838 | 91.92% | 173338731 | 68.30% | 54872025 |
| 397 | 94653775 | 93.44% | 176883243 | 62.80% | 65720019 |
| 398 | 94164370 | 93.32% | 175752605 | 61.90% | 66968050 |
| 399 | 88567436 | 93.30% | 165258210 | 69.10% | 51033967 |
| 400 | 98416526 | 92.21% | 181506660 | 61% | 70857795 |
| 401 | 95476259 | 92.08% | 175822231 | 74% | 45652965 |
| 402 | 92721577 | 93.59% | 173558028 | 66.90% | 57505064 |
| 403 | 92259150 | 91.10% | 168094982 | 66.80% | 55805579 |
| 404 | 88083362 | 93.76% | 165177022 | 58.10% | 69219073 |
| 405 | 92673640 | 92.23% | 170944821 | 68.20% | 54331377 |
| 406 | 91470878 | 90.71% | 165950945 | 68.30% | 52630329 |
| 407 | 92289829 | 91.24% | 168411040 | 66.50% | 56472297 |
| 408 | 88452199 | 92.23% | 163152205 | 63.10% | 60181547 |
| 409 | 94457069 | 91.78% | 173385603 | 69.20% | 53418541 |
| 410 | 90804922 | 90.60% | 164542909 | 59.40% | 66759518 |
| 411 | 92170656 | 92.31% | 170159210 | 58.80% | 70080399 |
| 412 | 88015234 | 90.63% | 159541716 | 67.40% | 52066946 |
| 413 | 90896266 | 92.02% | 167291419 | 66.50% | 55964132 |
| 414 | 95267470 | 93.90% | 178918725 | 63.10% | 66019900 |
| 415 | 90307683 | 89.91% | 162395740 | 75.20% | 40311703 |
| 416 | 89954030 | 93.51% | 168237445 | 72.70% | 45963463 |
| 417 | 92393129 | 90.77% | 167722925 | 58.60% | 69448174 |
| 418 | 96504239 | 92.38% | 178305273 | 54.50% | 81161684 |
| 419 | 94181054 | 92.24% | 173736160 | 53.60% | 80633299 |
| 420 | 88132332 | 92.81% | 163596957 | 55.20% | 73288089 |
| 421 | 92289521 | 93.42% | 172434278 | 56.30% | 75417880 |
| 422 | 93722558 | 91.93% | 172308930 | 59.10% | 70468747 |
| 423 | 91533009 | 90.67% | 165985429 | 72.30% | 45947853 |
| 424 | 98072046 | 92.20% | 180847605 | 66.80% | 59997635 |
| 425 | 94242577 | 88.97% | 167693401 | 61.30% | 64908370 |
| 426 | 90280373 | 91.21% | 164689198 | 66.60% | 54965722 |
| 427 | 93086902 | 94.13% | 175245153 | 62.80% | 65237940 |
| 428 | 88128911 | 92.82% | 163601591 | 63.40% | 59947062 |
| 429 | 93573424 | 93.78% | 175506311 | 62.50% | 65822250 |
| 430 | 93868423 | 93.42% | 175381316 | 57.60% | 74397340 |
| 431 | 98482895 | 90.88% | 179010692 | 73.80% | 46966176 |
| 432 | 93638980 | 93.02% | 174207479 | 64.10% | 62462076 |
| 433 | 89719416 | 93.27% | 167369730 | 65.70% | 57463722 |
| 434 | 95073085 | 91.95% | 174833170 | 61.70% | 66880236 |
| 435 | 89822530 | 92.97% | 167011901 | 67.30% | 54612076 |
| 436 | 91834032 | 92.67% | 170198511 | 61.20% | 66085430 |
| 437 | 92515385 | 89.52% | 165636384 | 71.80% | 46708002 |
| 438 | 81638563 | 93.65% | 152909707 | 55.20% | 68573636 |
| 439 | 100452737 | 93.09% | 187024703 | 54.60% | 84975617 |
| 440 | 96805206 | 92.74% | 179556773 | 65.90% | 61168508 |
| 441 | 98930632 | 93.51% | 185023729 | 58.70% | 76505368 |
| 442 | 96017904 | 92.77% | 178147884 | 58.20% | 74503587 |
| 443 | 90936815 | 93.73% | 170469363 | 58.90% | 70128609 |
| 444 | 89671949 | 92.11% | 165189706 | 63.80% | 59734038 |
| 445 | 93655808 | 93.56% | 175242283 | 56.20% | 76800194 |
| 446 | 92481706 | 93.39% | 172736906 | 60.80% | 67797725 |
| 447 | 92242548 | 93.29% | 172115150 | 58.20% | 71974570 |
| 448 | 98958108 | 92.46% | 183000445 | 54.90% | 82605208 |
| 449 | 89329760 | 93.54% | 167121757 | 57.40% | 71144290 |
| 450 | 92546113 | 92.53% | 171258949 | 52.70% | 80939003 |
| 451 | 95495914 | 90.44% | 172734328 | 64.70% | 60986570 |
| 452 | 90617866 | 91.86% | 166476607 | 56.70% | 72097969 |
| 453 | 85249239 | 92.77% | 158175937 | 50% | 79019520 |
| 454 | 96473089 | 93.21% | 179838851 | 61.30% | 69667130 |
| 455 | 87622388 | 93.39% | 163658186 | 52.40% | 77843675 |
| 456 | 106753538 | 95.40% | 203694475 | 53.80% | 94184243 |
| 457 | 97876344 | 94.93% | 185836410 | 59.10% | 75983609 |
| 458 | 96724752 | 94.96% | 183708439 | 56.70% | 79591673 |
| 459 | 98803703 | 94.37% | 186480153 | 56.10% | 81804328 |
| 460 | 93016321 | 94.98% | 176700272 | 54% | 81212759 |
| 461 | 92084819 | 94.83% | 174656278 | 55.80% | 77212005 |
| 462 | 95001560 | 93.67% | 177982126 | 57.20% | 76218237 |
| 463 | 106809646 | 94.65% | 202187866 | 57.80% | 85413716 |
| 464 | 87839255 | 94.12% | 165355226 | 52.80% | 77967579 |
| 465 | 105811413 | 94.38% | 199739189 | 56.70% | 86405329 |
| 466 | 95541031 | 91.59% | 175007190 | 69.10% | 54148777 |
| 467 | 100430849 | 94.49% | 189789449 | 58.90% | 77935513 |
| 468 | 105982368 | 93.82% | 198873360 | 56.20% | 87132432 |
| 469 | 101514895 | 94.94% | 192751591 | 50.30% | 95763249 |
| 470 | 94660040 | 89.61% | 169654972 | 59.10% | 69446557 |
| 471 | 101901254 | 95.58% | 194798665 | 54.90% | 87802945 |
| 472 | 94699721 | 94.57% | 179109617 | 57.20% | 76588848 |
| 473 | 105237557 | 94.78% | 199487211 | 54.10% | 91466671 |
| 474 | 95567120 | 96.24% | 183956252 | 62.70% | 68536981 |
| 475 | 94395254 | 96.17% | 181562730 | 58.70% | 74947279 |
| 476 | 98761952 | 94.66% | 186975979 | 58.10% | 78341010 |
| 477 | 92467802 | 94.89% | 175485793 | 59.10% | 71696131 |
| 478 | 96982315 | 94.40% | 183094897 | 61.40% | 70687779 |
| 479 | 101831651 | 94.93% | 193331574 | 53.20% | 90575634 |
| 480 | 97300773 | 95.01% | 184887565 | 52.60% | 87728858 |
| 481 | 96666603 | 93.43% | 180640088 | 63.40% | 66139023 |
| 482 | 102251855 | 95.72% | 195760752 | 58.30% | 81682880 |
| 483 | 97637176 | 94.63% | 184781648 | 52.90% | 86991385 |
| 484 | 101283273 | 93.99% | 190390008 | 57.20% | 81456991 |
| 485 | 101091302 | 94.91% | 191881678 | 57.90% | 80701975 |
| 486 | 93013111 | 94.11% | 175065722 | 59.80% | 70354519 |
| 487 | 105604071 | 94.43% | 199445496 | 54.20% | 91328980 |
| 488 | 93988400 | 93.25% | 175281809 | 64.30% | 62488580 |
| 489 | 99689678 | 94.93% | 189267116 | 59.30% | 77096563 |
| 490 | 103505998 | 95.05% | 196756837 | 58.80% | 80978414 |
| 491 | 97155437 | 94.88% | 184359341 | 61% | 71943927 |
| 492 | 97162311 | 94.64% | 183899317 | 64.20% | 65825221 |
| 493 | 97734107 | 94.45% | 184624265 | 62.60% | 69118740 |
| 494 | 97047216 | 94.80% | 183998443 | 63.20% | 67785009 |
| 495 | 97440239 | 94.48% | 184125148 | 54.40% | 84029016 |
| 496 | 94981444 | 94.62% | 179740485 | 62.30% | 67796232 |
| 497 | 90898018 | 93.98% | 170847926 | 62.10% | 64797984 |
| 498 | 98192837 | 95.43% | 187415265 | 57.50% | 79655358 |
| 499 | 94635900 | 95.09% | 179972994 | 57% | 77352751 |
| 500 | 96304744 | 95.18% | 183331357 | 58.10% | 76877869 |
| 501 | 99152855 | 94.85% | 188094606 | 52.80% | 88830603 |
| 502 | 87912768 | 94.13% | 165498166 | 57.90% | 69747262 |
| 503 | 102072863 | 94.93% | 193786918 | 58.80% | 79862675 |
| 504 | 95626828 | 94.70% | 181125533 | 62.10% | 68618652 |
| 505 | 95462159 | 95.32% | 181984022 | 58.90% | 74786308 |
| 506 | 86963075 | 94.99% | 165212291 | 59.70% | 66591725 |
| 507 | 102034874 | 94.81% | 193475179 | 62.70% | 72089086 |
| 508 | 93891414 | 94.86% | 178132723 | 61.10% | 69241995 |
| 509 | 95377432 | 94.81% | 180862233 | 61.40% | 69852609 |
| 510 | 105657361 | 94.56% | 199815540 | 59.40% | 81070642 |
| 511 | 94505366 | 95.15% | 179845898 | 54.60% | 81647583 |
| 512 | 93503827 | 95.19% | 178005941 | 92.80% | 12853785 |
| 513 | 97584136 | 94.64% | 184705043 | 55.40% | 82434795 |
| 514 | 94742267 | 94.40% | 178876936 | 57.80% | 75464728 |
| 515 | 96662924 | 95.12% | 183899328 | 60.60% | 72487446 |
| 516 | 95153939 | 94.63% | 180092578 | 60.40% | 71351472 |
| 517 | 94091898 | 93.90% | 176704699 | 62.10% | 66893054 |
| 518 | 92269025 | 94.15% | 173743946 | 59.40% | 70526796 |
| 519 | 94381366 | 94.39% | 178175304 | 56.20% | 77967554 |
| 520 | 95654141 | 95.06% | 181862423 | 58% | 76350924 |
| 521 | 97093342 | 94.96% | 184396026 | 58.60% | 76254633 |
| 522 | 92388585 | 95.13% | 175771350 | 58.80% | 72358164 |
| 523 | 92060426 | 94.69% | 174350333 | 58% | 73313254 |
| 524 | 109881129 | 94.29% | 207219855 | 57.20% | 88731020 |
| 525 | 101050070 | 95.68% | 193365348 | 59% | 79184265 |
| 526 | 96318962 | 94.25% | 181563940 | 58.90% | 74608502 |
| 527 | 91688583 | 94.76% | 173766923 | 58.80% | 71569344 |
| 528 | 98328743 | 95.19% | 187205952 | 58.30% | 78136105 |
| 529 | 121446366 | 94.15% | 228685610 | 57.70% | 96642845 |
| 530 | 100138016 | 94.31% | 188880646 | 61.80% | 72140808 |
| 531 | 94415088 | 95.10% | 179586762 | 57.30% | 76701379 |
| 532 | 102141157 | 95.18% | 194426179 | 55.30% | 86963987 |
| 533 | 97730240 | 95.01% | 185713182 | 54.70% | 84165544 |
| 534 | 96131064 | 95.57% | 183745808 | 55.20% | 82243616 |
| 535 | 95363612 | 94.02% | 179323841 | 59% | 73469130 |
| 536 | 102903390 | 94.13% | 193719440 | 61.20% | 75199767 |
| 537 | 100346986 | 92.46% | 185567738 | 62.10% | 70414077 |
| 538 | 100722120 | 94.73% | 190831813 | 57.70% | 80788804 |
| 539 | 95881211 | 94.30% | 180836733 | 60.70% | 71070593 |
| 540 | 95567678 | 95.28% | 182111290 | 52.60% | 86301224 |
| 541 | 97359400 | 95.05% | 185084691 | 59.30% | 75373328 |
| 542 | 97578199 | 94.91% | 185213389 | 61.30% | 71606892 |
| 543 | 96910187 | 95.58% | 185246359 | 53.70% | 85782395 |
| 544 | 112780954 | 95.18% | 214694984 | 52.20% | 102724183 |
| 545 | 93275663 | 95.35% | 177880631 | 56.20% | 77840263 |
| 546 | 83354267 | 94.64% | 157767241 | 50.80% | 77638649 |
| 547 | 90439586 | 95.09% | 171994479 | 59.60% | 69448972 |
| 548 | 103168200 | 93.69% | 193320598 | 52.20% | 92492657 |
| 549 | 98614058 | 95.08% | 187530774 | 52.90% | 88392266 |
| 550 | 98772349 | 94.43% | 186545174 | 51.80% | 89907393 |
| 551 | 96029438 | 91.99% | 176675593 | 58.20% | 73793839 |
| 552 | 94987817 | 94.13% | 178823254 | 55.70% | 79172902 |
| 553 | 98177356 | 95.20% | 186920472 | 52.80% | 88148676 |
| 554 | 94374784 | 94.06% | 177536940 | 56.20% | 77690572 |
| 555 | 99184295 | 94.78% | 188008498 | 61.80% | 71824464 |
| 556 | 96475347 | 93.23% | 179896780 | 56.40% | 78377785 |
| 557 | 95325137 | 94.82% | 180772579 | 55.80% | 79913858 |
| 558 | 99134429 | 96.11% | 190559341 | 62.80% | 70917976 |
| 559 | 95543580 | 94.65% | 180869530 | 57.70% | 76457595 |
| 560 | 101595656 | 95.06% | 193147151 | 55.40% | 86101711 |
| 561 | 91435512 | 94.70% | 173172445 | 55% | 77845967 |
| 562 | 95844954 | 93.06% | 178382933 | 60% | 71414095 |
| 563 | 93400224 | 94.81% | 177107821 | 55.50% | 78810835 |
| 564 | 97693345 | 93.78% | 183240742 | 55.80% | 81060064 |
| 565 | 111457841 | 94.01% | 209556407 | 54.40% | 95599806 |
| 566 | 100924314 | 94.69% | 191128298 | 60.80% | 74961152 |
| 567 | 95146106 | 94.99% | 180753766 | 60% | 72328773 |
| 568 | 92308177 | 94.52% | 174506357 | 60.40% | 69065020 |
| 569 | 93055956 | 94.71% | 176261034 | 53.60% | 81708334 |
| 570 | 94736443 | 94.54% | 179120753 | 59.50% | 72515103 |
| 571 | 101645086 | 94.82% | 192751968 | 59.50% | 78029324 |
| 572 | 95245554 | 93.82% | 178711513 | 58.70% | 73752412 |
| 573 | 94462242 | 94.09% | 177753492 | 58.60% | 73591688 |
| 574 | 94920698 | 94.50% | 179406360 | 57.90% | 75451736 |
| 575 | 93789701 | 95.31% | 178776881 | 57.80% | 75530134 |

**Supplemental Table 3:** Listings of Ensembl gene (ENSG) associations identified by negative binomial regression for each trait (one table per trait: Max OXPHOS, VO_2_ peak, 400-meter Walk Speed, Leg Strength, Thigh Muscle Mass, and Whole Body D3Cr).

| Gene_symbol | ensembl_gene_id | Base Mean | Log 2-Fold Change | Standard Error | Wald Statistic | Unadjusted p-value | FDR adjusted p-value |
| --- | --- | --- | --- | --- | --- | --- | --- |
| CAT | ENSG00000121691 | 3536.640566 | 0.020799112 | 0.01104257 | 1.88353845 | 0.05962743 | 0.083478408 |
| GPX1 | ENSG00000233276 | 1606.329124 | 0.029656353 | 0.01414079 | 2.09722027 | 0.03597408 | 0.053961121 |
| GPX2 | ENSG00000176153 | 0.92068084 | -0.165889668 | 0.09453044 | -1.754881 | 0.07927967 | 0.104054566 |
| GPX3 | ENSG00000211445 | 2841.022862 | -0.051201476 | 0.02124657 | -2.4098701 | 0.0159582 | 0.027926854 |
| GPX4 | ENSG00000167468 | 3526.885745 | 0.007563184 | 0.01457408 | 0.51894748 | 0.60379737 | 0.633987236 |
| GPX5 | ENSG00000116157 | 147.9612591 | -0.005490288 | 0.01434978 | -0.3826044 | 0.70201311 | 0.702013113 |
| GPX6 | ENSG00000164294 | 76.44044373 | -0.071198016 | 0.02335975 | -3.0478932 | 0.00230452 | 0.004399535 |
| GRX1 | ENSG00000173221 | 1037.515987 | -0.07024026 | 0.02151681 | -3.2644365 | 0.00109682 | 0.002559247 |
| GRX2 | ENSG00000023572 | 212.0634461 | 0.081154104 | 0.01309099 | 6.19923588 | 5.67E-10 | 3.97E-09 |
| GRX3 | ENSG00000108010 | 830.1114115 | 0.016501173 | 0.00705916 | 2.33755493 | 0.01941035 | 0.031355178 |
| PRX1 | ENSG00000117450 | 4508.929957 | -0.039831525 | 0.01098233 | -3.6268749 | 2.87E-04 | 8.61E-04 |
| PRX2 | ENSG00000167815 | 5205.327895 | 0.08403996 | 0.01156045 | 7.2696099 | 3.61E-13 | 7.57E-12 |
| PRX3 | ENSG00000165672 | 6278.727427 | 0.040946481 | 0.00981352 | 4.1724552 | 3.01E-05 | 1.05E-04 |
| PRX4 | ENSG00000123131 | 383.2419114 | -0.047625975 | 0.01341724 | -3.5496115 | 3.86E-04 | 0.001012725 |
| PRX5 | ENSG00000126432 | 3652.338857 | 0.068025145 | 0.01423265 | 4.77951467 | 1.76E-06 | 7.38E-06 |
| PRX6 | ENSG00000117592 | 8536.515726 | 0.028092468 | 0.00902325 | 3.11334166 | 0.00184982 | 0.003884617 |
| SOD1 | ENSG00000142168 | 3355.603106 | 0.006267947 | 0.01105577 | 0.56693895 | 0.57075566 | 0.630835204 |
| SOD2 | ENSG00000112096 | 17284.53832 | 0.08830396 | 0.01324324 | 6.66785309 | 2.60E-11 | 2.73E-10 |
| SOD3 | ENSG00000109610 | 154.289445 | -0.059715478 | 0.035917 | -1.6625966 | 0.09639322 | 0.119073977 |
| TRX1 | ENSG00000136810 | 623.353772 | -0.012914325 | 0.01628655 | -0.7929442 | 0.42781034 | 0.499112063 |
| TRX2 | ENSG00000100348 | 2592.598218 | 0.060850851 | 0.01082092 | 5.62344331 | 1.87E-08 | 9.83E-08 |

**Supplemental Table 4:** Listings of Ensembl gene (ENSG) associations identified by negative binomial regression for each trait (one table per trait: Max OXPHOS, VO_2_ peak, 400-meter Walk Speed, Leg Strength, Thigh Muscle Mass, and Whole Body D3Cr).

| Gene_symbol | ensembl_gene_id | Base Mean | Log 2-Fold Change | Standard Error | Wald Statistic | Unadjusted p-value | FDR adjusted p-value |
| --- | --- | --- | --- | --- | --- | --- | --- |
| CAT | ENSG00000121691 | 3592.269893 | 0.020388334 | 0.0165513 | 1.2318302 | 0.21801252 | 0.331462201 |
| GPX1 | ENSG00000233276 | 1646.479136 | 0.034821591 | 0.0209871 | 1.6591916 | 0.09707718 | 0.196342976 |
| GPX2 | ENSG00000176153 | 0.905649986 | -0.421707114 | 0.1450658 | -2.9070058 | 0.00364906 | 0.008514485 |
| GPX3 | ENSG00000211445 | 2853.575891 | -0.037429566 | 0.0319792 | -1.170435 | 0.24182594 | 0.338556315 |
| GPX4 | ENSG00000167468 | 3605.99008 | -0.012158441 | 0.0215882 | -0.5631988 | 0.57329951 | 0.668849423 |
| GPX5 | ENSG00000116157 | 150.5454421 | 0.030594153 | 0.0217993 | 1.4034494 | 0.16048287 | 0.280845028 |
| GPX6 | ENSG00000164294 | 76.54495587 | 0.016771646 | 0.0359288 | 0.4668027 | 0.64064108 | 0.708076981 |
| GRX1 | ENSG00000173221 | 1052.973874 | -0.100403095 | 0.033604 | -2.987828 | 0.00280968 | 0.0073754 |
| GRX2 | ENSG00000023572 | 214.7983405 | 0.070105428 | 0.0206891 | 3.3885232 | 7.03E-04 | 0.002951344 |
| GRX3 | ENSG00000108010 | 838.7854228 | 0.017615545 | 0.0107991 | 1.6312086 | 0.10284632 | 0.196342976 |
| PRX1 | ENSG00000117450 | 4591.513808 | -0.069009194 | 0.0165704 | -4.1646142 | 3.12E-05 | 2.91E-04 |
| PRX2 | ENSG00000167815 | 5329.397319 | 0.074544418 | 0.0181895 | 4.0982201 | 4.16E-05 | 2.91E-04 |
| PRX3 | ENSG00000165672 | 6378.991017 | 0.076555706 | 0.0145009 | 5.2793887 | 1.30E-07 | 2.72E-06 |
| PRX4 | ENSG00000123131 | 389.6228736 | -0.018189237 | 0.0205381 | -0.8856353 | 0.37581411 | 0.493256013 |
| PRX5 | ENSG00000126432 | 3742.08287 | 0.066436822 | 0.0216872 | 3.0634087 | 0.00218831 | 0.006564928 |
| PRX6 | ENSG00000117592 | 8688.82927 | 0.005530201 | 0.0139186 | 0.3973245 | 0.69112821 | 0.725684624 |
| SOD1 | ENSG00000142168 | 3417.991118 | -0.011146816 | 0.0167137 | -0.6669256 | 0.50481965 | 0.62360074 |
| SOD2 | ENSG00000112096 | 17516.63393 | 0.081735281 | 0.0204854 | 3.9899299 | 6.61E-05 | 3.47E-04 |
| SOD3 | ENSG00000109610 | 150.8857732 | -0.003270022 | 0.0510508 | -0.0640543 | 0.94892697 | 0.948926967 |
| TRX1 | ENSG00000136810 | 630.5753855 | -0.030223607 | 0.0246937 | -1.2239402 | 0.2209748 | 0.331462201 |
| TRX2 | ENSG00000100348 | 2650.274681 | 0.054658119 | 0.0168583 | 3.2422103 | 0.00118606 | 0.004151226 |

**Supplemental Table 5:** Listings of Ensembl gene (ENSG) associations identified by negative binomial regression for each trait (one table per trait: Max OXPHOS, VO_2_ peak, 400-meter Walk Speed, Leg Strength, Thigh Muscle Mass, and Whole Body D3Cr).

| Gene_symbol | ensembl_gene_id | Base Mean | Log 2-Fold Change | Standard Error | Wald Statistic | Unadjusted p-value | FDR adjusted p-value |
| --- | --- | --- | --- | --- | --- | --- | --- |
| CAT | ENSG00000121691 | 3585.390585 | 0.010189612 | 0.0113074 | 0.901147 | 0.3675101 | 0.615558922 |
| GPX1 | ENSG00000233276 | 1635.574598 | -0.016821335 | 0.0140978 | -1.1931929 | 0.2327938 | 0.464367537 |
| GPX2 | ENSG00000176153 | 0.920250442 | -0.10843793 | 0.0929261 | -1.1669263 | 0.2432401 | 0.464367537 |
| GPX3 | ENSG00000211445 | 2864.281901 | -0.075568556 | 0.021428 | -3.5266198 | 4.21E-04 | 0.004419457 |
| GPX4 | ENSG00000167468 | 3589.335283 | -0.004412526 | 0.0144678 | -0.3049889 | 0.7603746 | 0.887103668 |
| GPX5 | ENSG00000116157 | 149.6814679 | 0.012769492 | 0.014578 | 0.8759444 | 0.3810603 | 0.615558922 |
| GPX6 | ENSG00000164294 | 76.71538891 | -0.033008035 | 0.0238003 | -1.3868757 | 0.1654797 | 0.406290641 |
| GRX1 | ENSG00000173221 | 1047.642848 | -0.052082612 | 0.0222875 | -2.3368576 | 0.0194466 | 0.06806306 |
| GRX2 | ENSG00000023572 | 213.1942236 | 8.68E-04 | 0.0139702 | 0.062138 | 0.9504529 | 0.997975568 |
| GRX3 | ENSG00000108010 | 834.8093103 | 0.003685894 | 0.0071914 | 0.5125403 | 0.6082729 | 0.798358176 |
| PRX1 | ENSG00000117450 | 4578.206837 | -0.051338745 | 0.0108619 | -4.7265132 | 2.28E-06 | 4.80E-05 |
| PRX2 | ENSG00000167815 | 5286.351421 | 0.001869263 | 0.0122899 | 0.1520974 | 0.8791101 | 0.971648035 |
| PRX3 | ENSG00000165672 | 6327.809818 | 0.03145825 | 0.0101224 | 3.1077866 | 0.0018849 | 0.009895942 |
| PRX4 | ENSG00000123131 | 388.0849099 | 0.008715982 | 0.0138445 | 0.6295623 | 0.528981 | 0.740573403 |
| PRX5 | ENSG00000126432 | 3713.455874 | 0.005632956 | 0.0145095 | 0.3882245 | 0.6978499 | 0.862049876 |
| PRX6 | ENSG00000117592 | 8636.675112 | -0.005852851 | 0.0092106 | -0.6354496 | 0.5251353 | 0.740573403 |
| SOD1 | ENSG00000142168 | 3401.027127 | -0.021963822 | 0.010992 | -1.9981722 | 0.045698 | 0.137093988 |
| SOD2 | ENSG00000112096 | 17352.42193 | 0.018829422 | 0.0138546 | 1.3590695 | 0.1741246 | 0.406290641 |
| SOD3 | ENSG00000109610 | 155.8195125 | -0.112999695 | 0.0356863 | -3.1664728 | 0.001543 | 0.009895942 |
| TRX1 | ENSG00000136810 | 629.0138476 | -0.041550626 | 0.0163342 | -2.5437756 | 0.0109662 | 0.046057839 |
| TRX2 | ENSG00000100348 | 2629.257654 | -9.63E-06 | 0.0112872 | -8.53E-04 | 0.9993192 | 0.999319217 |

**Supplemental Table 6:** Listings of Ensembl gene (ENSG) associations identified by negative binomial regression for each trait (one table per trait: Max OXPHOS, VO_2_ peak, 400-meter Walk Speed, Leg Strength, Thigh Muscle Mass, and Whole Body D3Cr).

| Gene_symbol | ensembl_gene_id | Base Mean | Log 2-Fold Change | Standard Error | Wald Statistic | Unadjusted p-value | FDR adjusted p-value |
| --- | --- | --- | --- | --- | --- | --- | --- |
| CAT | ENSG00000121691 | 3571.291718 | 0.011211287 | 0.0138526 | 0.8093286 | 0.4183262 | 0.627489228 |
| GPX1 | ENSG00000233276 | 1638.93964 | 0.009448744 | 0.017391 | 0.5433136 | 0.5869139 | 0.821521591 |
| GPX2 | ENSG00000176153 | 0.914530847 | -0.056704787 | 0.1163233 | -0.487476 | 0.6259212 | 0.821521591 |
| GPX3 | ENSG00000211445 | 2851.242963 | -0.033795366 | 0.0263004 | -1.284976 | 0.1988007 | 0.417481539 |
| GPX4 | ENSG00000167468 | 3598.765541 | -0.001204347 | 0.0177946 | -0.06768 | 0.9460401 | 0.993342067 |
| GPX5 | ENSG00000116157 | 149.5434931 | 0.033227187 | 0.0174603 | 1.9030088 | 0.0570394 | 0.19703381 |
| GPX6 | ENSG00000164294 | 76.45041 | -0.006446824 | 0.0293871 | -0.219376 | 0.8263574 | 0.913342381 |
| GRX1 | ENSG00000173221 | 1042.001154 | 0.026690029 | 0.0272123 | 0.980808 | 0.3266874 | 0.527725843 |
| GRX2 | ENSG00000023572 | 213.113651 | 0.03830537 | 0.0169269 | 2.2629841 | 0.0236367 | 0.101144012 |
| GRX3 | ENSG00000108010 | 834.6912074 | 0.014764507 | 0.0086455 | 1.7077726 | 0.0876785 | 0.230156184 |
| PRX1 | ENSG00000117450 | 4567.218891 | -0.054772574 | 0.0133599 | -4.099765 | 4.14E-05 | 8.68E-04 |
| PRX2 | ENSG00000167815 | 5291.612745 | 0.016514817 | 0.0151116 | 1.0928554 | 0.2744573 | 0.480300288 |
| PRX3 | ENSG00000165672 | 6332.472884 | 0.028042082 | 0.012431 | 2.25582 | 0.0240819 | 0.101144012 |
| PRX4 | ENSG00000123131 | 388.0841437 | 0.01988785 | 0.0168595 | 1.179626 | 0.238149 | 0.454648079 |
| PRX5 | ENSG00000126432 | 3717.657427 | 0.032855621 | 0.0178503 | 1.8406154 | 0.0656779 | 0.19703381 |
| PRX6 | ENSG00000117592 | 8618.505492 | -0.014643164 | 0.0112878 | -1.297253 | 0.194544 | 0.417481539 |
| SOD1 | ENSG00000142168 | 3393.928407 | -0.003649322 | 0.0134542 | -0.271239 | 0.7862069 | 0.913342381 |
| SOD2 | ENSG00000112096 | 17357.4281 | 0.040939991 | 0.0168627 | 2.4278361 | 0.0151892 | 0.101144012 |
| SOD3 | ENSG00000109610 | 155.5398227 | -0.155010779 | 0.0438706 | -3.533363 | 4.10E-04 | 0.004308239 |
| TRX1 | ENSG00000136810 | 627.2693115 | -1.45E-04 | 0.02002 | -0.007233 | 0.9942293 | 0.994229312 |
| TRX2 | ENSG00000100348 | 2632.277607 | 0.004273069 | 0.0138585 | 0.3083347 | 0.7578276 | 0.913342381 |

**Supplemental Table 7:** Listings of Ensembl gene (ENSG) associations identified by negative binomial regression for each trait (one table per trait: Max OXPHOS, VO_2_ peak, 400-meter Walk Speed, Leg Strength, Thigh Muscle Mass, and Whole Body D3Cr).

| Gene_symbol | ensembl_gene_id | Base Mean | Log 2-Fold Change | Standard Error | Wald Statistic | Unadjusted p-value | FDR adjusted p-value |
| --- | --- | --- | --- | --- | --- | --- | --- |
| CAT | ENSG00000121691 | 3591.448278 | -0.016333568 | 0.02539747 | -0.643118 | 0.52014757 | 0.606838826 |
| GPX1 | ENSG00000233276 | 1635.286662 | 0.011275682 | 0.031605 | 0.35676895 | 0.72126478 | 0.75732802 |
| GPX2 | ENSG00000176153 | 0.923960722 | -0.368989151 | 0.21121477 | -1.7469855 | 0.08063985 | 0.182093355 |
| GPX3 | ENSG00000211445 | 2874.879088 | -0.045383919 | 0.04805962 | -0.9443253 | 0.34500344 | 0.517505167 |
| GPX4 | ENSG00000167468 | 3589.282415 | -0.024173591 | 0.03239402 | -0.7462362 | 0.45552472 | 0.562707001 |
| GPX5 | ENSG00000116157 | 149.6223278 | 6.48E-04 | 0.03263767 | 0.01985046 | 0.98416267 | 0.984162668 |
| GPX6 | ENSG00000164294 | 76.89552489 | 0.08815068 | 0.05285874 | 1.66766507 | 0.09538223 | 0.182093355 |
| GRX1 | ENSG00000173221 | 1047.924975 | -0.024251191 | 0.04914783 | -0.4934336 | 0.62170625 | 0.687149013 |
| GRX2 | ENSG00000023572 | 213.3906652 | 0.127321044 | 0.03054245 | 4.16865785 | 3.06E-05 | 3.22E-04 |
| GRX3 | ENSG00000108010 | 834.384473 | 0.026404406 | 0.01566926 | 1.68510889 | 0.09196756 | 0.182093355 |
| PRX1 | ENSG00000117450 | 4579.187885 | -0.128075093 | 0.02403429 | -5.3288489 | 9.88E-08 | 2.08E-06 |
| PRX2 | ENSG00000167815 | 5294.492168 | 0.049649165 | 0.02731575 | 1.81760194 | 0.06912499 | 0.182093355 |
| PRX3 | ENSG00000165672 | 6330.557468 | 0.068630913 | 0.02250904 | 3.0490377 | 0.00229576 | 0.012052723 |
| PRX4 | ENSG00000123131 | 387.7326614 | 0.023251376 | 0.03015147 | 0.77115221 | 0.44061672 | 0.562707001 |
| PRX5 | ENSG00000126432 | 3713.956045 | 0.055678424 | 0.03232615 | 1.7223956 | 0.08499788 | 0.182093355 |
| PRX6 | ENSG00000117592 | 8663.968863 | -0.024263245 | 0.020413 | -1.1886173 | 0.2345903 | 0.410533027 |
| SOD1 | ENSG00000142168 | 3400.837399 | -0.048465734 | 0.02422776 | -2.0004215 | 0.04545477 | 0.159091696 |
| SOD2 | ENSG00000112096 | 17371.52306 | 0.121637594 | 0.03061064 | 3.97370249 | 7.08E-05 | 4.95E-04 |
| SOD3 | ENSG00000109610 | 156.8308598 | -0.179850569 | 0.08014791 | -2.2439833 | 0.02483348 | 0.104300606 |
| TRX1 | ENSG00000136810 | 628.6504202 | -0.030828476 | 0.03593097 | -0.8579916 | 0.39089708 | 0.547255912 |
| TRX2 | ENSG00000100348 | 2633.3436 | 0.025874756 | 0.02501905 | 1.03420204 | 0.30104172 | 0.486298162 |

**Supplemental Table 8:** Listings of Ensembl gene (ENSG) associations identified by negative binomial regression for each trait (one table per trait: Max OXPHOS, VO_2_ peak, 400-meter Walk Speed, Leg Strength, Thigh Muscle Mass, and Whole Body D3Cr).

| Gene_symbol | ensembl_gene_id | Base Mean | Log 2-Fold Change | Standard Error | Wald Statistic | Unadjusted p-value | FDR adjusted p-value |
| --- | --- | --- | --- | --- | --- | --- | --- |
| CAT | ENSG00000121691 | 3557.926407 | -0.030349068 | 0.015417624 | -1.96846597 | 0.049014448 | 0.17446875 |
| GPX1 | ENSG00000233276 | 1628.663082 | 0.019402342 | 0.019293055 | 1.005664569 | 0.314576952 | 0.508162769 |
| GPX2 | ENSG00000176153 | 0.915597597 | 0.349529335 | 0.124353879 | 2.810763432 | 0.004942411 | 0.035261374 |
| GPX3 | ENSG00000211445 | 2865.490425 | 0.040180434 | 0.02967695 | 1.353927333 | 0.175759562 | 0.369095081 |
| GPX4 | ENSG00000167468 | 3579.788156 | 0.020945854 | 0.019938515 | 1.050522298 | 0.293478045 | 0.508162769 |
| GPX5 | ENSG00000116157 | 148.6481679 | 0.012547368 | 0.019947818 | 0.629009554 | 0.529342802 | 0.653894049 |
| GPX6 | ENSG00000164294 | 76.22789715 | 0.087978097 | 0.03245723 | 2.710585543 | 0.006716452 | 0.035261374 |
| GRX1 | ENSG00000173221 | 1041.145766 | 0.046023684 | 0.030569217 | 1.505556518 | 0.132181058 | 0.308422468 |
| GRX2 | ENSG00000023572 | 211.548143 | 0.070257016 | 0.018477007 | 3.802402404 | 1.43E-04 | 0.003009294 |
| GRX3 | ENSG00000108010 | 828.9073641 | 0.019206444 | 0.00979289 | 1.961264172 | 0.049848214 | 0.17446875 |
| PRX1 | ENSG00000117450 | 4549.437954 | -0.012668234 | 0.015280099 | -0.82906758 | 0.407066165 | 0.610599248 |
| PRX2 | ENSG00000167815 | 5271.862115 | -8.83E-04 | 0.01672705 | -0.05280418 | 0.957887929 | 0.957887929 |
| PRX3 | ENSG00000165672 | 6282.480784 | 0.01692661 | 0.014036435 | 1.205905168 | 0.227854058 | 0.43499411 |
| PRX4 | ENSG00000123131 | 385.6266566 | 0.035039922 | 0.01883315 | 1.86054489 | 0.062808473 | 0.18842542 |
| PRX5 | ENSG00000126432 | 3702.216113 | 0.012815591 | 0.019996753 | 0.640883625 | 0.521598295 | 0.653894049 |
| PRX6 | ENSG00000117592 | 8586.790786 | -0.004999731 | 0.012627298 | -0.39594625 | 0.692144678 | 0.765002012 |
| SOD1 | ENSG00000142168 | 3382.217714 | 0.003149757 | 0.015199144 | 0.207232498 | 0.835828288 | 0.877619703 |
| SOD2 | ENSG00000112096 | 17215.38143 | 0.009041537 | 0.019004453 | 0.475758883 | 0.634246166 | 0.73995386 |
| SOD3 | ENSG00000109610 | 156.0650668 | -0.086321405 | 0.049453829 | -1.74549485 | 0.080898768 | 0.212359267 |
| TRX1 | ENSG00000136810 | 623.8251393 | 0.060872991 | 0.022349346 | 2.723703491 | 0.006455443 | 0.035261374 |
| TRX2 | ENSG00000100348 | 2618.571945 | -0.010610119 | 0.01546764 | -0.68595593 | 0.492740902 | 0.653894049 |

## Notes

### Competing Interest Statement

S.R. Cummings is a consultant to Bioage Labs. P.M. Cawthon is a consultant to and owns stock in MyoCorps. All other authors declare no conflict of interest.

### Author Declarations

Western Institutional Review Board Copernicus Group (WCG IRB 20180764) gave ethical approval for this work.

